# Evaluating parameter selection and analysis approaches on quality and reproducibility of functional MRS

**DOI:** 10.1101/2025.05.08.25327171

**Authors:** Duanghathai Pasanta, David J. Lythgoe, Nicolaas A. Puts

## Abstract

Functional magnetic resonance spectroscopy (fMRS) extends conventional MRS by acquiring data while participants receive stimuli or are engaged in a task, with analysis focused on segmenting data to align with stimulus- or task-related metabolite changes. In this study, we propose and systematically evaluate three fMRS analysis pipelines: block, event-related, and sliding window approaches, to optimise parameter selection and assess reproducibility with respect to data quality. Using empirical and simulated fMRS datasets, we examine the impact of data quality parameters and investigate the influence of the number of transients per block/window on the trade-off between reproducibility and temporal resolution with respect to data quality. Our results show that while the number of transients required for quantification can be reduced for Glx quantification in both block and event-related analyses, a minimum of 32 transients is required for reliable GABA+ measurement using spectral editing sequences. Optimal data quality, characterized by low noise and a spectral linewidth of 6-8 Hz, is preferred, especially for analyses with fewer transients. These findings highlight the importance of balancing data quality and acquisition parameters to ensure robust fMRS outcomes. Additionally, they provide a framework and consideration for implementing high temporal resolution analysis of GABA and glutamate, positioning fMRS as a powerful tool for advancing our understanding of neural mechanisms underlying brain function in both health and disease.

## Introduction

Functional magnetic resonance spectroscopy (fMRS) is an emerging technique for investigating dynamic changes in brain neurometabolites. Compared to conventional MRS, which provides an average measurement of metabolite levels across a number of transients, fMRS acquires data during task or stimulus processing, capturing transient neurochemical responses offering valuable insights into neurochemical processes underlying brain function. fMRS analyses differ from static MRS in their requirements for higher temporal resolution to track the time-course of task-related changes in metabolite levels. This involves segmenting the MRS data in a meaningful way to align with the stimulus- or task-related metabolite response. This however presents unique challenges in both experimental design and analysis in balancing temporal resolution and reliability of quantification, where higher temporal resolution often means averaging across a smaller number of transients and thus, lower signal to noise ratio (SNR).

Most fMRS studies to date have adopted designs analogous to those of fMRI, which are block and/or event-related paradigms, based on stimulus presentation and analysis. Block designs typically use consecutive transients for metabolite quantification, event-related designs assign the transients based on stimulus presentation. Studies suggest that different types of paradigms, or rather, different temporal resolutions, assess different processes within the brain. For example, block designs reflect averaged changes over time^1,2^, while event-related designs offer stimulus-related changes. This is supported by meta-analyses of fMRS studies showing a higher effect size for GABA and Glx for event-related designs^1,3^. However, the optimal number of transients required for reliable metabolite quantification to detect changes in metabolite levels during fMRS remains a subject of debate. Finally, as does the hemodynamic response function, the metabolite response likely has a temporal lag (the degree of which remains unknown) and thus sliding window approaches are also common.

Current consensus recommendations for conventional MRS parameters at 3 T propose the use of 240 transients with a 27 ml voxel when performing spectral editing for GABA^4,5^. Other studies challenge this consensus, suggesting that a lower number of transients, 18 transients for Glx and 32 transients for GABA+, may suffice^6,7^. This approach by using lower number of transient for metabolite quantification, however, comes with caveats, as it requires high-quality data reflected by good shimming, minimal participant movement, and a high SNR ratio. In fMRS studies, the most common ways to increase temporal resolution involve reducing number of transients, either by acquiring fewer transients during scan or by segmenting the data in smaller chunks for spectral fitting. Given that SNR scales with the square root of the number of transients, reducing number of transients, whether during acquisition or during spectral fitting, should have similar effects on quantification error. However, this raises critical questions about the optimal trade-offs between temporal resolution, data quality, and the number of transients required for metabolite quantification in fMRS experiments.

To address this, we evaluated three analysis pipelines: block, event-related, and sliding window analyses, using both existing and simulated datasets. The primary goal of this study was to examine how varying the numbers of transients in each analysis method affected the measurement of metabolite levels in fMRS data under specific data quality conditions. Our hypothesis is that, if the metabolite levels in MRS data remain relatively stable in the absence of a task, reducing the number of transients for quantification should not systematically alter the *measured* metabolite levels compared to the baseline measured with the full number of transients. We simulated fMRS data with known changes in GABA+ (GABA+macromolecules) and Glx (glutamate+glutamine) levels to mimic fMRS data acquired with MEGA-PRESS, with simulated data varying across levels of data quality.

Using these simulated fMRS data, we investigated the effects of both the fewer number of transients during spectral fitting and data quality on the reliability of high-temporal resolution analysis of fMRS. We hypothesised that the number of transients and data quality would influence the reliability of metabolite quantification. Specifically, we predicted that high data quality would enable the reliable detection of metabolite changes, while poor data quality would increase susceptibility to quantification error and potentially obscure expected, simulated metabolite changes.

## Methods

### fMRS Data

#### Baseline fMRS data

Previously acquired MEGA-PRESS MRS data from the “Big GABA” dataset ^8^ were used as a testbed for the proposed analyses. The Big GABA dataset included baseline data from 25 scan sites at 3 T. Thus, we expect levels to remain stable. All participants were healthy adults. These data are freely available online (https://www.nitrc.org/projects/biggaba/).

In this analysis, we used 20 datasets from each MRI manufacturer to replicate the typical participant numbers seen in (f)MRS studies, resulting in a total of 60 datasets. These datasets came from Philips sites (P1 and P3, n=20), GE sites (G5 and G7, n=20), and Siemens sites (S1 and S3, n=20). Common scan parameters for MEGA-PRESS data acquisitions across sites were 320 transients (160 ON and 160 OFF transients; 10 min and 40 seconds acquisition, TE/TR 68/2000ms, with a 3x3x3 cm^3^ voxel placed in the medial parietal lobe. Example averaged spectra from each site included in this study are shown in Supplementary Figure 1.

#### Simulated fMRS data

For simulated data, we simulated MEGA-PRESS fMRS spectra of 14 metabolites using FID-A ^9^ with the following parameters of B_0_ = 3 T, TE = 68 ms, TR = 2000 ms, 2000 Hz spectral width, and 2048 data points. The metabolites were simulated for MEGA-PRESS with hard localisation pulses, using the FID-A function *run_simMegaPressShaped_fast*^10^. The editing pulse shape is single banded gaussian editing pulse, with duration of 3.5 ms. The simulated metabolites included Aspartate (Asp), Creatine (Cr), Phosphocreatine (PCr), Glycerophosphocholine (GPC), Phosphocholine (PCh), GABA, Glutamine (Gln), glutamate (Glu), Glutathione (GSH), Myo-inositol (Ins), Lactate (Lac), N-Acetylaspartate (NAA), N-Acetylaspartylglutamate (NAAG), Scyllo-inositol (Scyllo), and macromolecules (MM), with estimated average concentrations and lineshapes based on previous studies (Supplementary Table 1)^11–13^. The MM peaks were simulated using Gaussian basis functions with a signal intensity in the multiplier of one proton, based on previous studies^14^ (Supplementary Table 2) where the co-edited MM underlying the GABA peak at 3.0 ppm was simulated with a simple spin system of two protons that coupled with the MM peak at 1.7 ppm with a linewidth of 14 Hz^15^. Residual water peak was modelled as a singlet with a Gaussian function at

#### 4.68 ppm and a concentration of 300 mM

The simulated metabolite spectra, which scale individual metabolites to estimated *in-vivo* concentrations by using *op_ampScale* function, were then combined to create baseline edit-ON and edit-OFF MEGA-PRESS spectra without exogenous noise and scan imperfections. Subsequently, MEGA-PRESS time series were generated by interleaving between edit-ON and edit-OFF spectra, during which GABA and glutamate were modulated to mimic stimulus-evoked changes in metabolite levels in fMRS. We implemented a stimulus block design, alternating blocks without metabolite modulation (RESTs) with blocks with metabolite modulations (FUNCs) into REST1-FUNC1-REST2-FUNC2 (Figure 2A) similar to many fMRS studies^1,3,16^. Each block comprises 80 transients, totalling 320 transients for the entire simulated spectrum. Within each fMRS block, stimuli were applied in an interleaved manner, starting with stim-ON for two transients (4s) followed by stim-OFF for two transients (4s), continuing until the end of each FUNC block. During FUNC blocks, GABA levels decreased by 20% and glutamate levels increased by 20% from baseline where the changes were instantaneous. While these may be large, this is based on a previous study that showed a 20% decrease in GABA+ and a 20% increase in Glx in response to tactile stimulation^17^. The resulting simulated fMRS spectra were saved in FID-A .mat format.

The simulation also incorporated scan imperfections to emulate real spectra, including a total linear frequency drift of 2 Hz and a negative phase drift of 1 degree over the whole scan to mimic subtle frequency drift in scanner found in a previous study^18^, using the FID-A *op_makeFreqDrift* function. To evaluate the impact of spectral quality on data fitting, we introduced three standard deviation levels of normally distributed random noise (noise_SD_) of 3SD, 5SD, and 10SD, and three line broadening factors of 6 Hz (excellent), 8 Hz (good), and 10 Hz (acceptable). The line broadening was performed by applying exponential apodisation on the simulated MEGA-PRESS time series with the *op_filter* function. A total of 20 spectra were simulated for each noise/LW level (Figure 1). All spectra and fitted models are plotted in Supplementary Figure 2.

**Figure 1.**
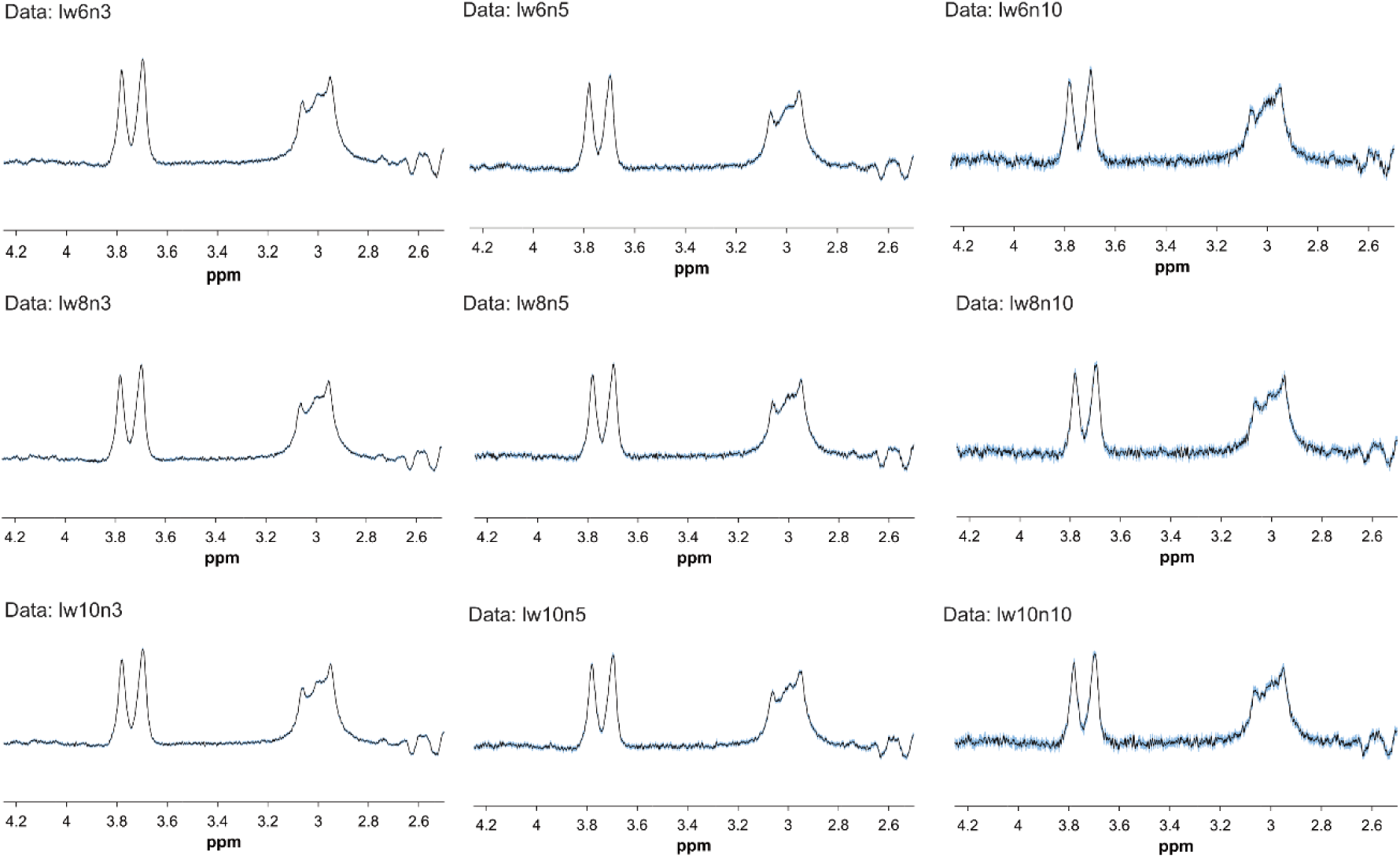
Averaged simulated MEGA-PRESS spectra with 95%CI shaded in blue. lw=linewidth(Hz); n=noise_SD_.

### fMRS analysis

All data, both baseline and simulated, were analysed using MATLAB^19^ with an in-house adaptation of the Gannet software^20^; a package tailored to the analysis of edited MRS. Three MRS spectroscopy analysis methods were implemented: block, event-related, and sliding window analysis for both thebaseline and the simulated data.

#### fMRS analysis of baseline data

The block analysis approach was performed by binning each acquisition into sub-difference spectra with different bin sizes (16, 32, 64, and 128 transients). For sliding window analysis, we chose window widths of 16, 32, 64, and 80 transients with a sliding step of 4 transients. The choice of window widths is a multiple of 8, as phase cycling is typically applied in blocks of 2, 4, 6, 8, or 16 transients^21,22^. The metabolites were quantified in ratio to total creatine (tCr) and in institutional units (i.u.).

#### fMRS analysis of simulated data

All data were processed through the same analysis pipeline, using an in-house modification of Gannet version 3.3.1 to load spectra simulated with FID-A. The same analysis pipeline within Gannet, that was used in the previous section on the baseline BIG-GABA data analyses was used, where the transients were segmented based on the type of analysis approach, that is block analysis, event-related analysis and sliding window analysis.

In the block analysis, the simulated fMRS data were quantified for metabolite levels based on the rest and fMRS blocks, each comprising 80 transients (FUNC1 and FUNC2) (Figure 2B). Event-related analysis was analysed based on how the stimuli were presented within each block. To allow for sufficient SNR, we segmented transients corresponding to the stimulus-ON and stimulus-OFF periods within each FUNC block and grouping them into two groups of 20 transients each to maintain adequate SNR while capturing the stimulus-locked changes during these periods. For each FUNC block, this resulted in two stim-ON phases of 20 transients and two stim-OFF phases of 20 transients (FUNC1 into ON1, ON2, OFF1, OFF2; FUNC2 into ON3, ON4, OFF3 and OFF4) (Figure 2C). Finally, sliding window analysis was applied to track the temporal changes of metabolite levels, using a window width of 40 transients and a step size of four transients of interleaving edit-ONs and edit-OFFs transients assuming two step phase cycling^22^ (Figure 2D).

**Figure 2.**
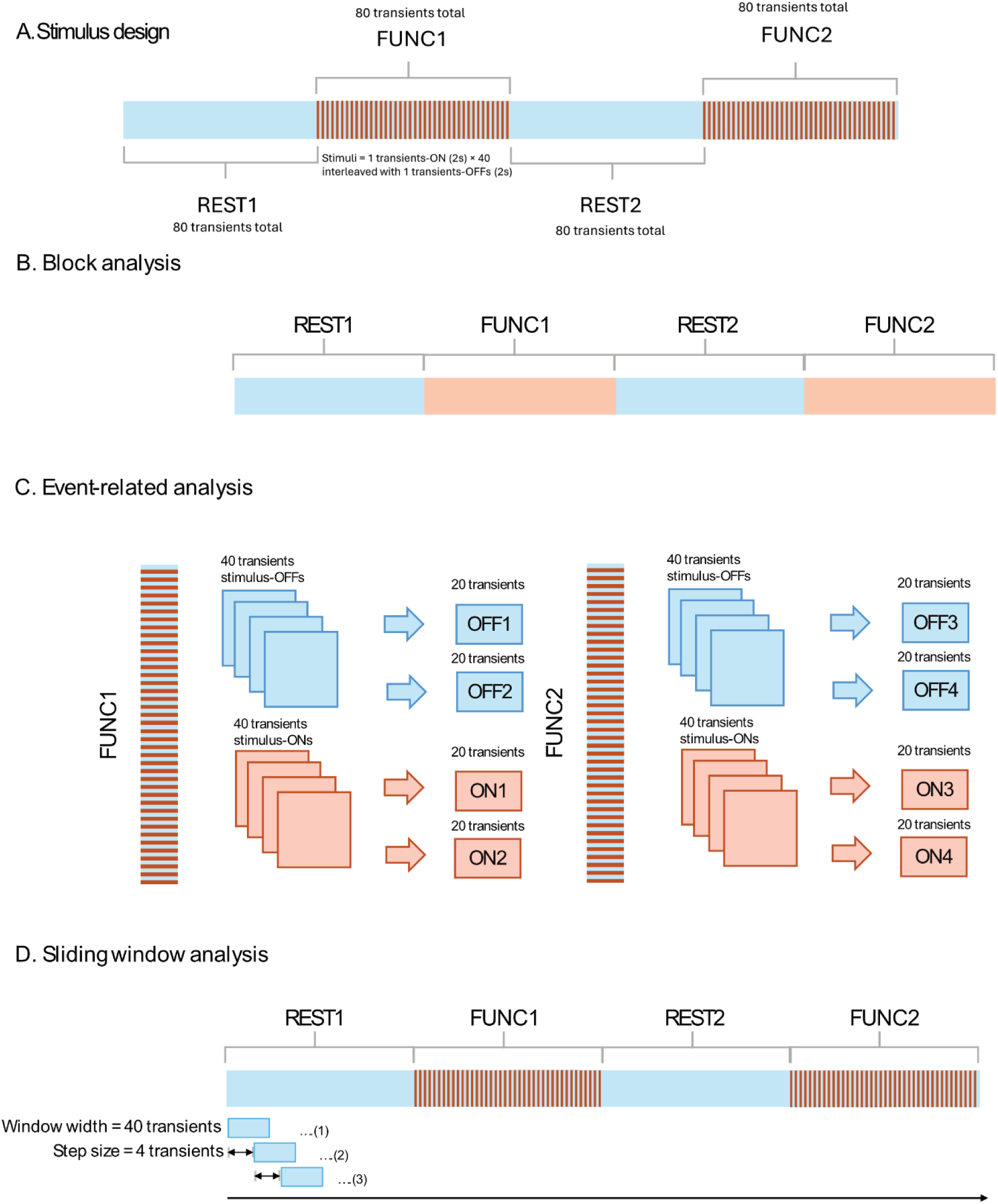
Schematic representations of the stimulus applied to simulated MEGA-PRESS fMRS and analysis approaches. (A) Schematic representation of the stimulus, (B) Block analysis approach, (C) Event-related analysis approach, (D) Sliding window analysis approach.

### Statistical analysis

All statistical analyses were performed in R version 4.2.3. To ensure MRS data quality, both visual and quantitative assessments of quality metrics were undertaken to exclude data affected by artefacts and outliers. After visual inspection for artefacts, subsequent data quality control criteria were applied sequentially. The first two criteria were related to spectral quality: data were excluded if the FWHM of NAA ≥ 10 Hz or %fit error exceeded 50%. These values were chosen to identify extreme erroneous values, based on the previous Big GABA study at 24 research sites that showed average %fit error of ∼9% and average NAA linewidth of ∼8 Hz^23^. The third criterion was to exclude the fitted concentrations if the target metabolites’ (GABA+/tCr, Glx/tCr, GABA/i.u., and Glx/i.u.) deviated from the median by more than 2.5 times the median absolute deviation (MAD). The same exclusion criteria were applied to all simulated spectra to mirror the quality assurance procedures conducted on *in vivo* spectra, irrespective of spectrum quality.

A Shapiro-Wilk test was used to assess the normality of the data distribution. To evaluate whether metabolite quantification from a reduced number of transients (i.e., static and sliding window analysis) aligned with metabolite levels derived from the full complement of 320 transients, correlation coefficients were calculated using the Spearman correlation test. Interclass correlations were not used in this current study due to a singular fit warning and models failed to converge which this suggests that variance of the random intercepts estimated to be 0.

A Kruskal-Wallis test was then applied to the results of the block analyses to determine if there were any significant changes in metabolite levels across the different numbers of windows used. Note that given this was data acquired at baseline, we did not expect any significant differences.

### Bayesian analysis

We then conducted additional Bayesian analyses on both block and sliding window fitting approach to evaluate the impact of the number of transients on metabolite levels on both baseline MRS and simulated fMRS data. The motivation for using Bayesian statistics was its flexibility in fitting the observed data. This includes incorporating an autoregressive term to account for autocorrelation between data points in sliding window analyses and enabling quantitative testing for evidence of no effect (null hypothesis)^24^. We implemented a Bayesian linear mixed model, followed by a subsequent test of practical equivalence using the *brms* and *bayestestR* packages in R^25,26^. Weakly informative priors were applied across all Bayesian analyses.

Bayesian linear mixed effects model for baseline fMRS data

We fitted a Bayesian linear mixed model to predict the effects of the number of transients on metabolite levels, accounting for variability at both the individual and site levels:

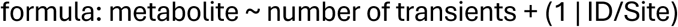

For the sliding window analysis, we introduced an additional autoregressive term to account for autocorrelation between data points, a consequence of the sliding window analysis method:

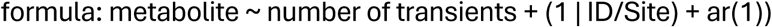

Bayesian linear mixed effects model for simulated fMRS data

For simulated data, the metabolite concentration values from REST1 were used as the baseline/reference for all models. Weakly informative priors were applied across all Bayesian analyses. For the block and event-related analyses, we adopted the same model used for the BIG-GABA analysis in the previous section to predict the impact of stimulus blocks on metabolite levels, accounting for within-subject variation. The formula for this model is:

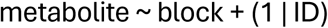

For the sliding window analysis of simulated data, we introduced an additional autoregressive term to address the autocorrelation inherent in the data points in similar manner to model used in baseline data. The classification of blocks (i.e., REST1, FUNC1, REST2, FUNC2) was based on the time of the middle of the window for each metabolite acquired. The formula for this model is:

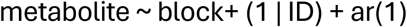

To avoid erroneously rejecting the null hypothesis of no effect, we then performed a test for practical equivalence following Bayesian linear mixed effects modelling. This test can be used to assess whether an observed effect is meaningfully different from zero, or another specific value within the context-defined "null-hypothesis" or range indicating a negligible effect size, also known as Range of Practical Equivalence (ROPE)^27,28^. The method involves checking if the 95% Highest Density Interval (HDI) of the posterior distributions falls within the ROPE. I the 95% HDI lie entirely outside the ROPE, the effect is deemed practically significant. Conversely, if the 95% HDI is fully within the ROPE, the effect is considered practically equivalent to null, implying the effect or difference is negligible. When the 95% HDI overlaps with the ROPE, the result is inconclusive, indicating that more data may be needed to make a definitive assessment.

After model fitting, the test for practical equivalence, using the same criteria as in the baseline fMRS data analysis was also applied here to determine whether the fMRS block is associated with a change in metabolite levels. If the 95% HDI fully fell within the ROPE region (-0.1 to 0.1 times the standard deviation of the outcome variable^28^), the effect was considered practically equivalent to null.

## Results

### Baseline fMRS data

#### Block analysis of baseline data

The quality metric of the MRS data included in the block analysis is reported in Table 1.

**Table 1.**
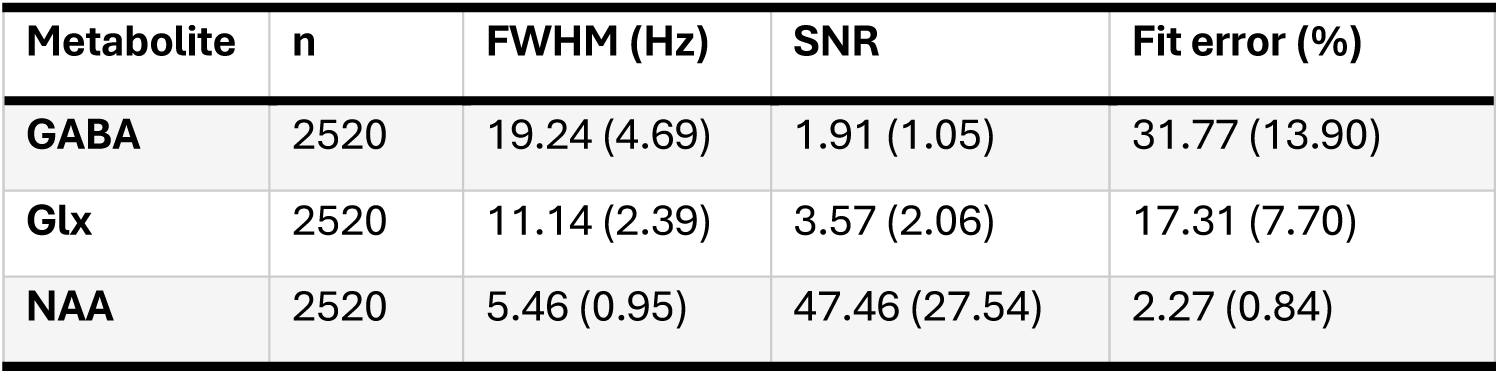
quality metric of MRS data included in the analysis regardless of the number of transients. The values are presented as mean (SD).

The block analyses demonstrate a tighter data distribution with an increasing number of transients for both GABA+ and Glx, regardless of the units of measurement (Cr, i.u.) (Figure 3), as has been shown before^29–32^. A Kruskal-Wallis test revealed no significant differences between GABA+ and Glx across the bins utilised in the block analysis for both units of measurement, suggesting stability across the acquisition.

**Figure 3.**
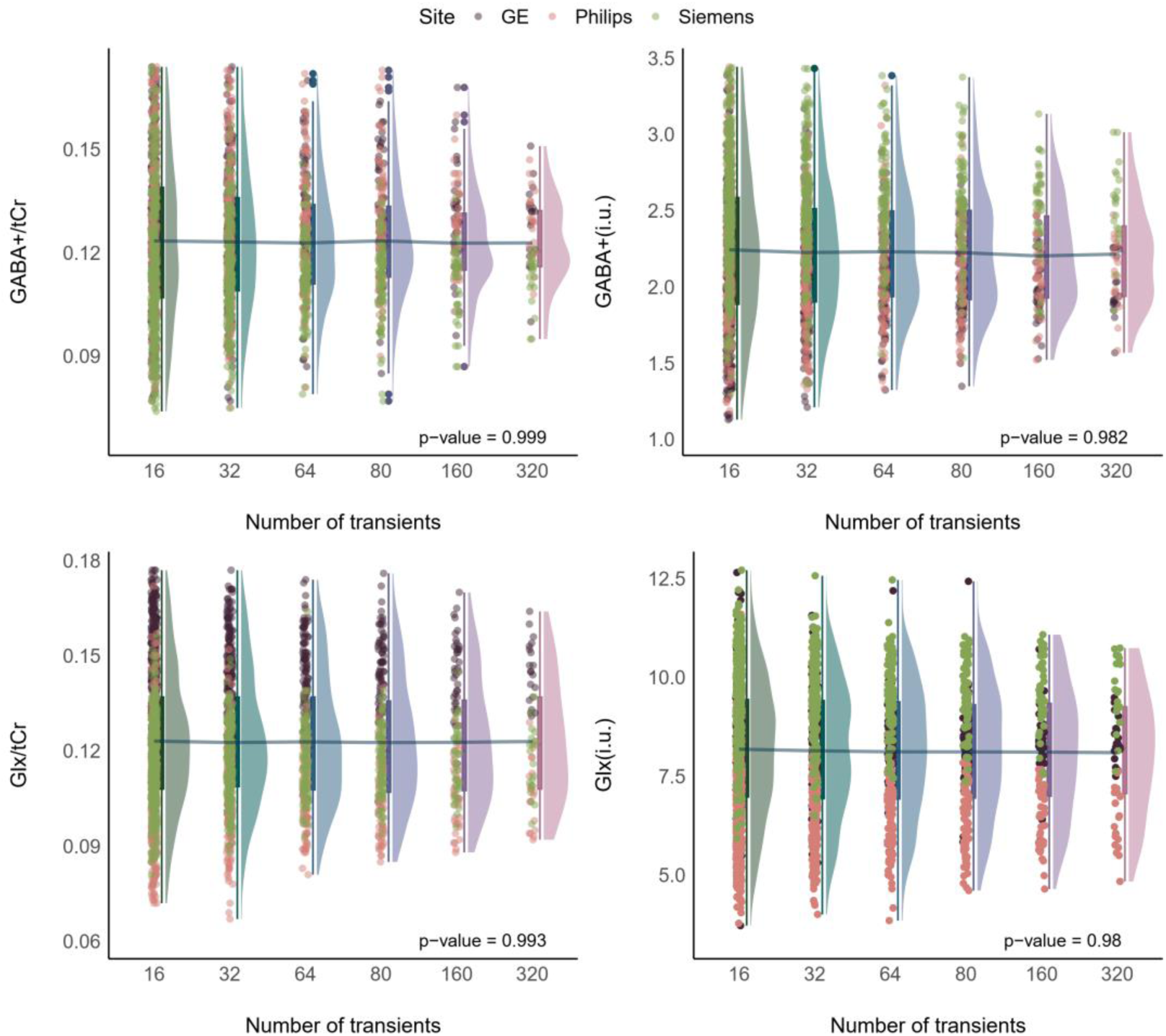
Metabolite levels for GABA+/tCr, GABA+/i.u., Glx/tCr, and Glx/i.u. across block analyses employing varying numbers of transients (16, 32, 64, 80, 160, 320), with 320 being the total number of transients available for each spectrum. P-values are from the Kruskal-Wallis test. The blue line across plots connects the mean metabolite values acquired from the number of transients used.

Figure 4 shows a correlation map between GABA+/tCr and Glx/tCr results from binned quantifications (transients = 16, 32, 64, 80, 160) and the reference values of GABA+/tCr and Glx/tCr, which were obtained using the full set of transients (transients = 320). Overall, Spearman correlations reveal a consistent trend for both GABA+/tCr and Glx/tCr, with the Spearman correlation coefficient increases with number of transients used in the analysis. For Glx/tCr, Spearman correlation coefficients indicate strong consistency with the reference, ranging from a minimum of 0.58 at 16 transients to a maximum of 0.93 at 160 transients. Conversely, for GABA+/tCr, visual inspection of the Spearman correlation coefficients indicates weaker consistency across the transient range, with coefficients varying from 0.02 at 16 transients to 0.72 at 160 transients.

**Figure 4.**
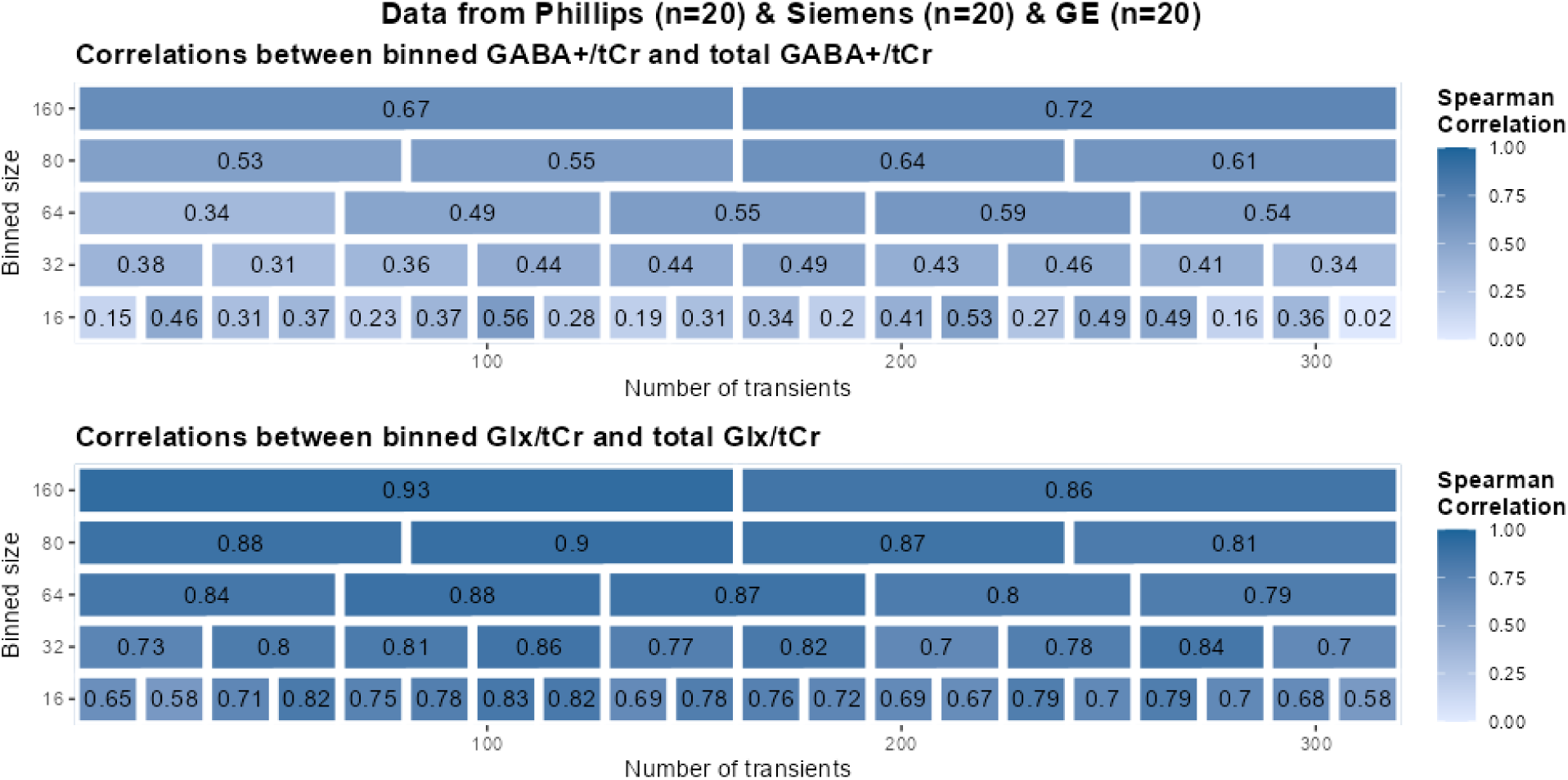
Pearson correlation map comparing metabolite levels from binned quantifications (transients = 16, 32, 64, 80, 160) with those obtained using the complete set of transients (transients = 320) across various sites, for GABA+/tCr and Glx/tCr.

Interestingly, the correlation map for GABA+ and Glx in institutional units (i.u.) reveals a marginal improvement in correlation coefficients between metabolites obtained from block analysis compared to reference concentrations, especially notable for GABA+ (i.u.) (Figure 5). In the case of Glx (i.u.), robust correlations were observed, with coefficients ranging from a minimum of 0.74 at 16 transients to a maximum of 0.94 at 160 transients. For GABA+ (i.u.), there was a visible increase in correlation coefficients compared to GABA+/tCr, with coefficients varying from 0.3 at 16 transients to 0.77 at 160 transients.

**Figure 5.**
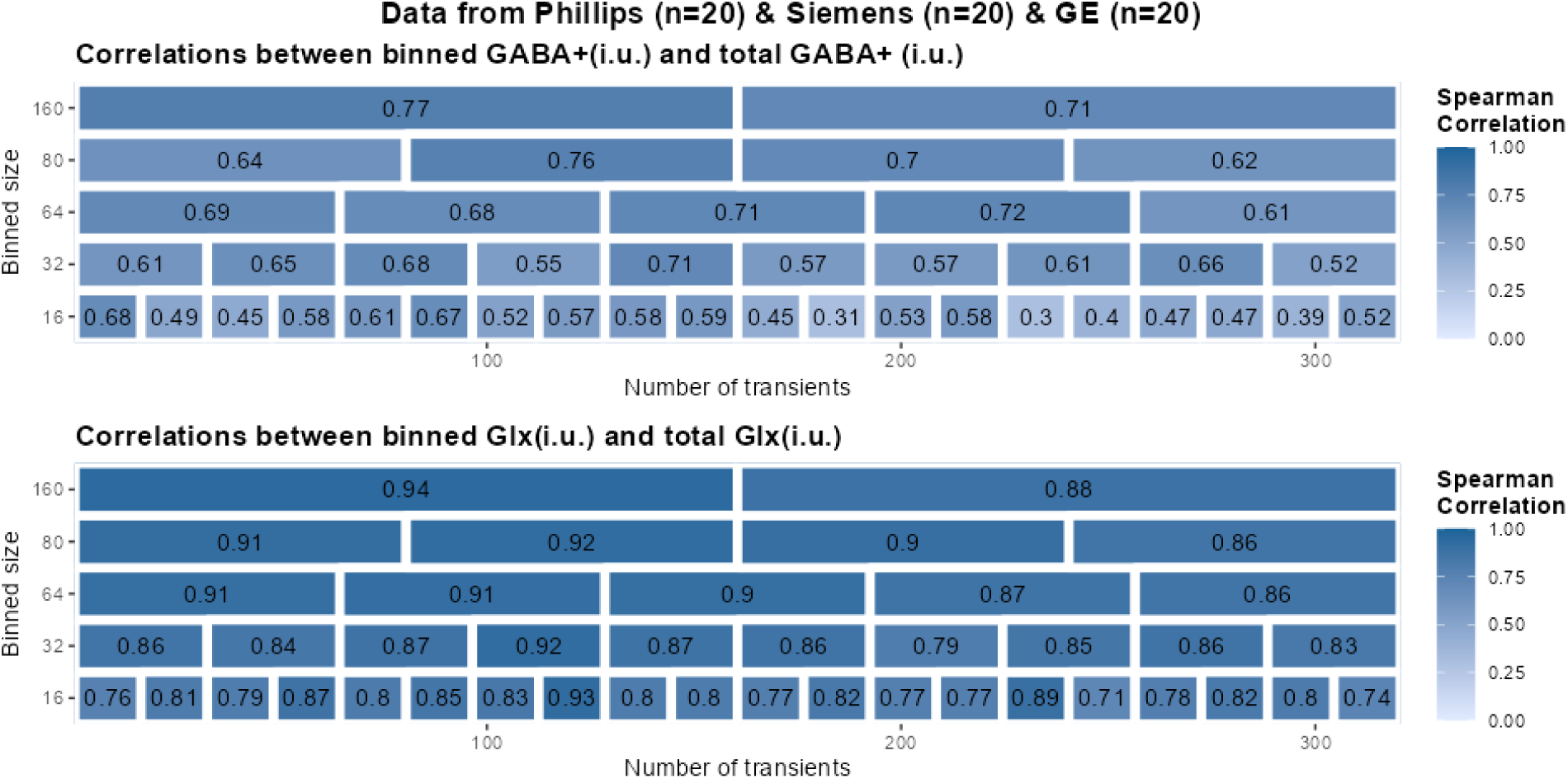
Pearson correlation map comparing metabolite levels from binned quantifications (transients = 16, 32, 64, 80, 160) with those obtained using the complete set of transients (transients = 320), across various sites, for GABA+(i.u.) and Glx(i.u.).

The results from Bayesian linear mixed model to assess the effect of the number of transients on metabolite levels quantified with block analyses are presented in Supplementary Table 3. The results generally indicate that varying the number of transients has a minimal practical impact on the quantification of GABA+ and Glx levels with block analysis for both units of i.u. and in ratio to tCr, with most coefficients demonstrating low probabilities of having a statistically significant effect on metabolite level.

These findings do, however, show that while concentrations correlate, there is some noise in the probable directionality of positive or negative changes in Glx across a baseline scan from the mixed directionality at different number of transients. Interestingly, the results showed mixed directional probabilities for both GABA+/tCr and GABA+ (i.u.). Blocks with sixteen transients showed the highest directional probability being 76.13% for GABA+/tCr and 83.78% for GABA+ (i.u.). Positive directional effects suggest that using a smaller number of transients consistently overestimates the measured concentration compared to the full acquisition, whereas negative directional effects suggest underestimation. There is around a 60% probability of being significant for both GABA+/tCr and GABA+ (i.u.), yet with low probabilities of large effects. This points to a minimal practical impact of the number of transients on GABA+ quantification in block analysis.

The outcomes from the test of practical equivalence demonstrate that a predominant portion of the 95% HDI falls within the ROPE for both GABA+ and Glx, irrespective of the units (Figure 6). Compared to Glx, GABA+ exhibits a slightly smaller percentage of the 95% HDI within the ROPE. Although the entirety of the 95% HDI does not fall within the ROPE, these findings imply a probable lack of practical effect regardless of the number of transients on the quantification of metabolite levels. What is important to note, however, is that with reduced numbers of transients, there is a higher likelihood of the HDI not falling within the ROPE, especially for GABA+. This is an important finding, particularly since these data were acquired at rest, and thus, we should not expect any effects or changes in metabolite concentration unless this reflects natural state changes which we will mention in the discussion.

**Figure 6.**
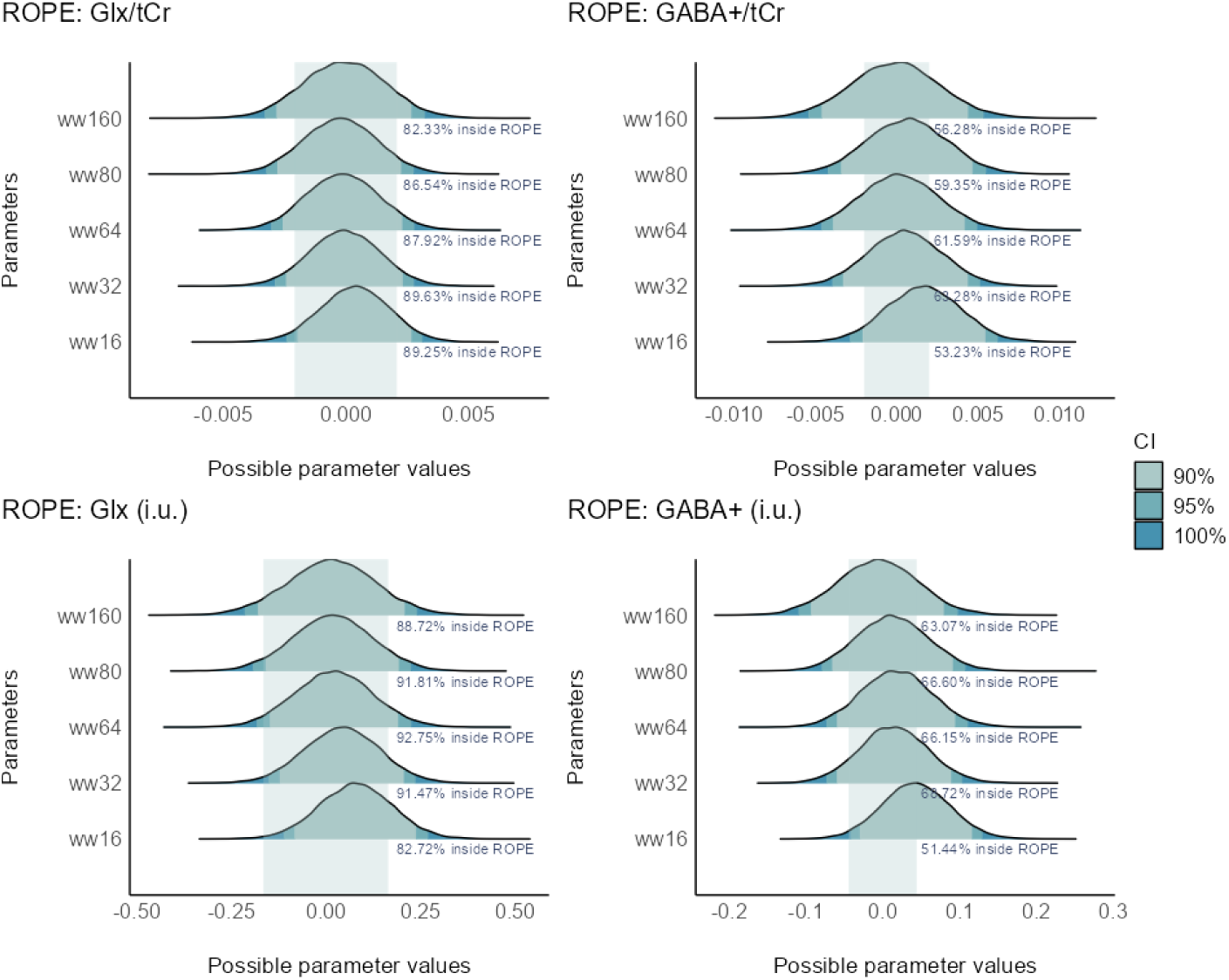
Posterior distribution plots of metabolite levels of baseline MRS data for varying transient numbers in quantification. Different shades represent the range within the posterior distribution that contains a specified proportion (90%, 95% and 100%) of the probability density. The superimposed green boxes show ROPE region (-0.1 to 0.1 times the standard deviation of the outcome variable ^28^ that denotes the range considered practically equivalent to a negligible effect. The percentage displayed shows the portion of the 95% HDI of the posterior distributions that falls within the ROPE.

### Sliding window analysis of baseline data

Figure 7 shows the metabolite levels acquired with varying transient sizes using sliding window analysis. Based on the visual inspection, variability of both GABA+ and Glx is lowest when a higher number of transients used in the analysis, with variability increasing when using a lower number of transients.

**Figure 7.**
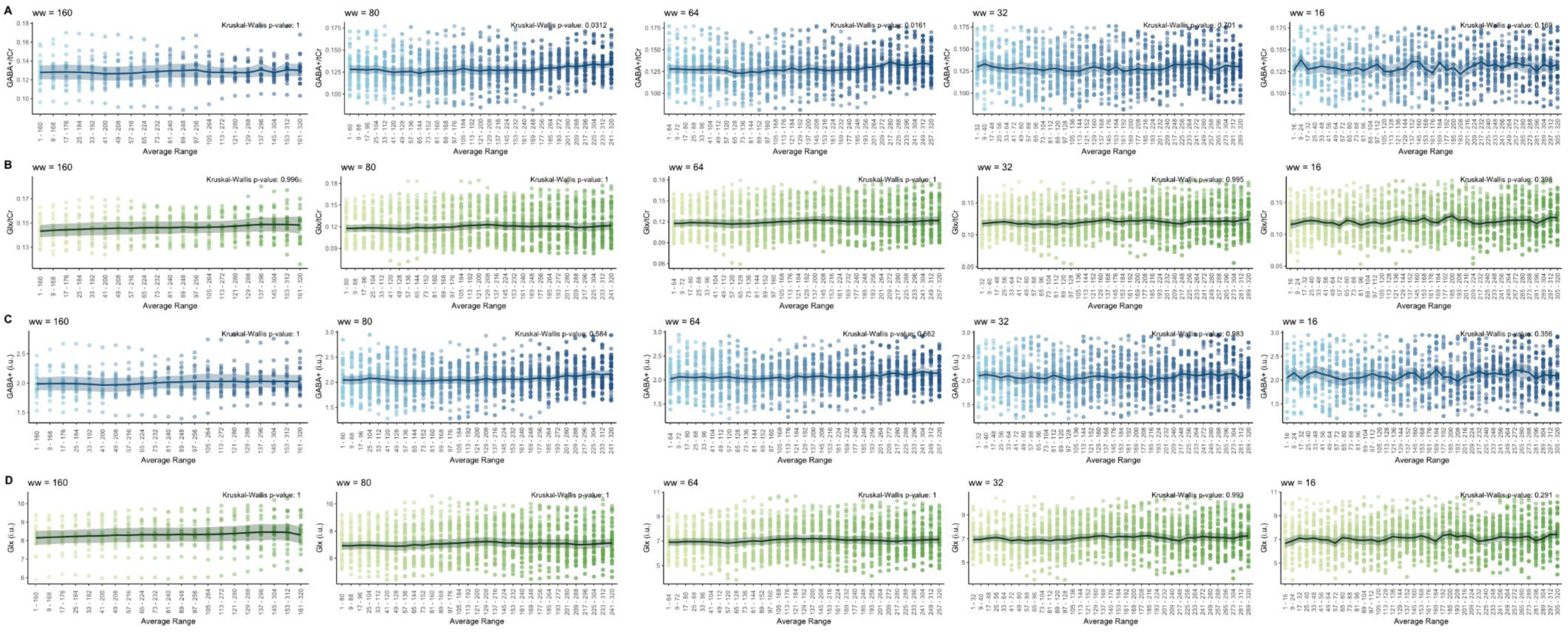
Metabolite levels relative to tCr and in i.u. unit, acquired using sliding window analysis with varying transient sizes (ww). (A) GABA+/tCr, (B) Glx/tCr, (C) GABA+(i.u.) and (D) Glx(i.u.). The solid line represents the mean metabolite level, while the shaded area indicates the 95% confidence interval for each window.

For GABA+/tCr and GABA+ (i.u.), the high probability of direction (pd) across all transient levels (pd = 0.92–1 for GABA+/tCr and pd = 0.92–1) suggests a probable existing positive effect of the number of transients on metabolite levels. As the transient size decreases, from 160 to 16, the effect and the probability of effect being significant and large increase. Particularly, 16 transients showed the highest impact, nearly 100% probability of a significant effect for both GABA+/tCr and GABA+ (i.u.) levels. However, the probability of the effects being large was more pronounced in GABA+/tCr, where the probability of being large was 99.94%, for GABA+/tCr and 64.89% for GABA+ (i.u.) at 16 transients.

These significant effects at 16 transients for both GABA+/tCr and GABA+ (i.u.) are visualised in the ROPE analysis is shown in Figure 8 where the 95% HDI lies entirely outside of the ROPE. This positive relationship between number of transients and GABA+ levels, suggests that a decrease in the number of transients in the sliding window analysis might lead to a false-positive increase in quantified GABA+/tCr and GABA+ (i.u.) levels.

**Figure 8.**
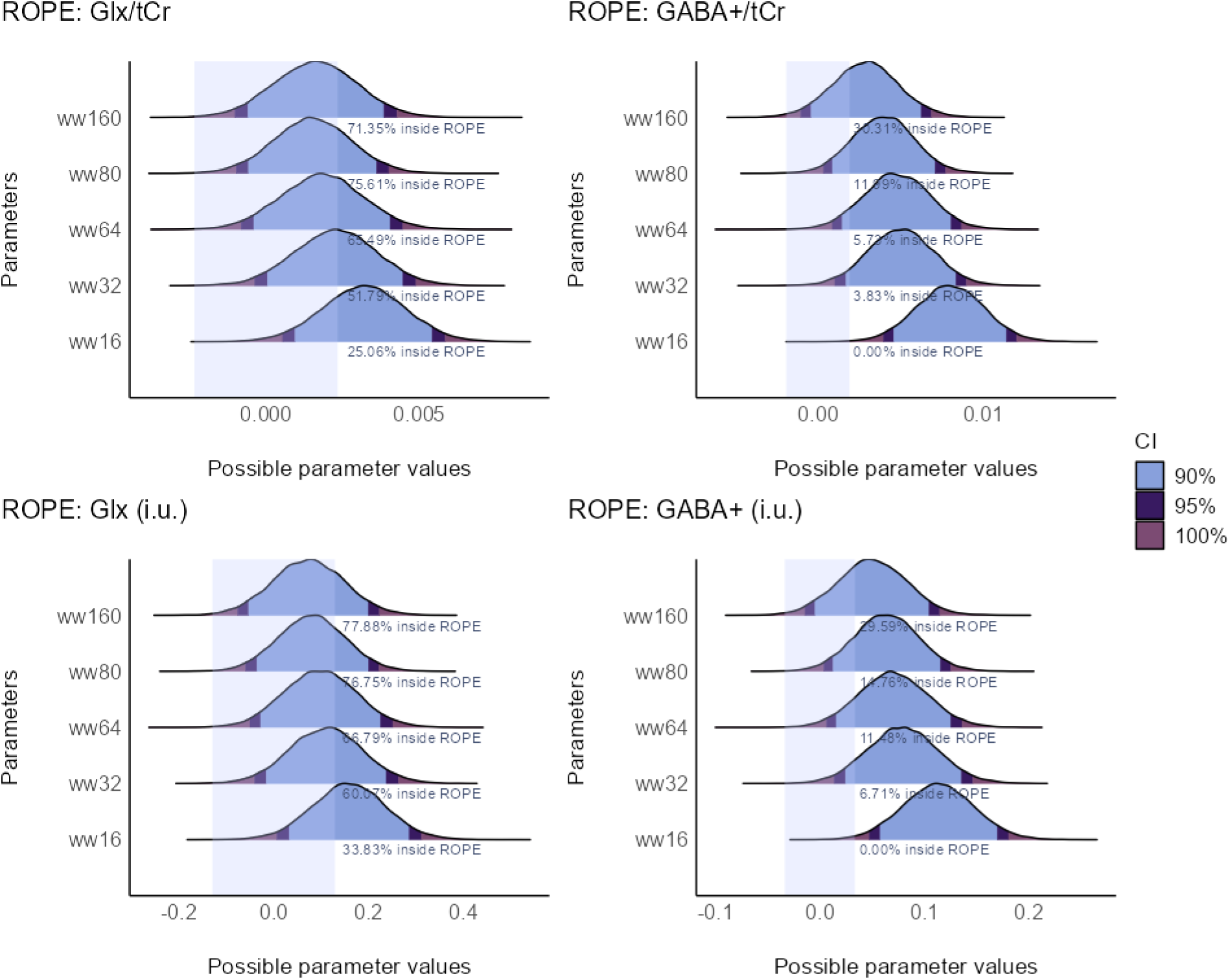
Posterior distribution plots of metabolite levels of baseline MRS data for varying transient numbers in quantification. Different shades represent the range within the posterior distribution that contains a specified proportion (90%, 95% and 100%) of the probability density. The superimposed purple boxes show ROPE region (-0.1 to 0.1 times the standard deviation of the outcome variable ^28^ that denotes the range considered practically equivalent to a negligible effect. The percentage displayed shows the portion of the 95% HDI of the posterior distributions that falls within the ROPE.

On the other hand, the effect of the number of transients on Glx/tCr and Glx (i.u.) showed varying degrees of significance and a generally modest effect size (Supplementary Table 4). Results showed pd in the range of 83.23%–99.05% directional probability of the effect size being positive across both Glx/tCr and Glx (i.u.), with a trend of increasing probability as transient size reduced. This result suggests the effect of the number transients being likely positive for Glx (i.u.) at 16 transients 95.26% (pd =97.94%) and certainly positive for Glx/tCr at 16 transients (pd=99.05%). Similarly with the block analysis counterpart, while probability of the effects being significant increases with reduced transient sizes (probability between 0.55–0.89), the effects are overall not considered large by the thresholds set in the analysis.

This suggests that while there was a trend towards higher Glx levels with smaller transients, the magnitude of this effect may not be substantial in all cases. These results are also visually shown in Figure 8, where a lower percentage of 95%HDI are within ROPE with reducing number of transients. However, a substantial portion of them is within ROPE, suggesting an inconclusive effect. When comparing Glx/tCr and Glx (i.u.), the probabilities of being positive, significant, and large vary, with generally higher probabilities of significance in the Glx/tCr analysis. This suggests a more consistent and clear influence of transient sizes on the ratio of Glx to creatine compared to Glx concentration alone.

### Simulated fMRS data

#### Block analysis of simulated fMRS data

The quality metrics of the simulated fMRS data included in the analysis after the removal of outliers and unsatisfactory data, are shown in *Table 2*.

**Table 2.** quality metrics of simulated fMRS data included in the block analysis for each quality level. lw=line width (Hz), n= noise_SD_. The data presents as mean (SD).

| Data | Metabolite | n | FWHM (Hz) | SNR | Fit Error (%) |
| --- | --- | --- | --- | --- | --- |
| **lw6n3** | Glx | 80 | 8.75 (0.25) | 7.22 (0.34) | 8.48 (0.57) |
|  | GABA | 80 | 17.5 (0.47) | 5.48 (0.25) | 11.17 (0.69) |
|  | NAA | 80 | 4.26 (0.02) | 106.81 (3.91) | 1.65 (0.04) |
| **lw6n5** | Glx | 80 | 8.61 (0.48) | 4.37 (0.31) | 12.16 (0.98) |
|  | GABA | 80 | 17.42 (0.85) | 3.27 (0.17) | 16.23 (0.98) |
|  | NAA | 80 | 4.26 (0.03) | 64.09 (2.75) | 1.76 (0.07) |
| **lw6n10** | Glx | 69 | 8.77 (0.95) | 2.24 (0.23) | 21.63 (2.34) |
|  | GABA | 58 | 17.18 (1.4) | 1.65 (0.15) | 29.21 (2.92) |
|  | NAA | 80 | 4.27 (0.07) | 31.95 (1.35) | 2.23 (0.12) |
| **lw8n3** | Glx | 80 | 8.69 (0.25) | 7.18 (0.35) | 8.48 (0.6) |
|  | GABA | 80 | 17.48 (0.54) | 5.40 (0.28) | 11.27 (0.74) |
|  | NAA | 80 | 4.26 (0.02) | 106.07 (4.23) | 1.64 (0.04) |
| **lw8n5** | Glx | 80 | 8.63 (0.54) | 4.35 (0.24) | 11.93 (0.96) |
|  | GABA | 78 | 17.28 (0.91) | 3.27 (0.19) | 15.9 (1.18) |
|  | NAA | 80 | 4.26 (0.03) | 64.06 (3.32) | 1.75 (0.07) |
| **lw8n10** | Glx | 71 | 8.62 (0.78) | 2.24 (0.21) | 21.56 (2.15) |
|  | GABA | 60 | 17.33 (1.41) | 1.64 (0.15) | 29.41 (2.95) |
|  | NAA | 80 | 4.26 (0.06) | 32.25 (1.45) | 2.2 (0.11) |
| **lw10n3** | Glx | 80 | 8.66 (0.32) | 7.18 (0.37) | 8.57 (0.54) |
|  | GABA | 80 | 17.4 (0.6) | 5.42 (0.28) | 11.36 (0.66) |
|  | NAA | 80 | 4.26 (0.02) | 106.8 (5.13) | 1.65 (0.05) |
| **lw10n5** | Glx | 79 | 8.64 (0.48) | 4.38 (0.25) | 12.00 (0.95) |
|  | GABA | 77 | 17.53 (0.85) | 3.3 (0.18) | 15.89 (1.09) |
|  | NAA | 80 | 4.26 (0.03) | 64.07 (2.84) | 1.76 (0.07) |
| **lw10n10** | Glx | 68 | 8.65 (0.91) | 2.24 (0.22) | 21.5 (2.46) |
|  | GABA | 65 | 17.13 (1.29) | 1.65 (0.12) | 29.03 (2.49) |
|  | NAA | 80 | 4.25 (0.06) | 32.27 (1.34) | 2.18 (0.12) |

Based on visual inspection, the expected reductions in GABA+ levels for both the ratio to tCr (Figure 9A) and the concentrations in i.u. (Supplementary Figure 3A) are lower with poorer data quality, especially with higher standard deviations of noise. On the other hand, the impact of LW on GABA+ change during fMRS blocks appears to be less noticeable when compared across the same noise level. The same trend in data quality affecting the results was observed for Glx/tCr (Figure 9B) and Glx (i.u.) (Supplementary Figure 3B). The expected increase in Glx levels during fMRS was less visible in datasets with higher noise_SD_.

**Figure 9.**
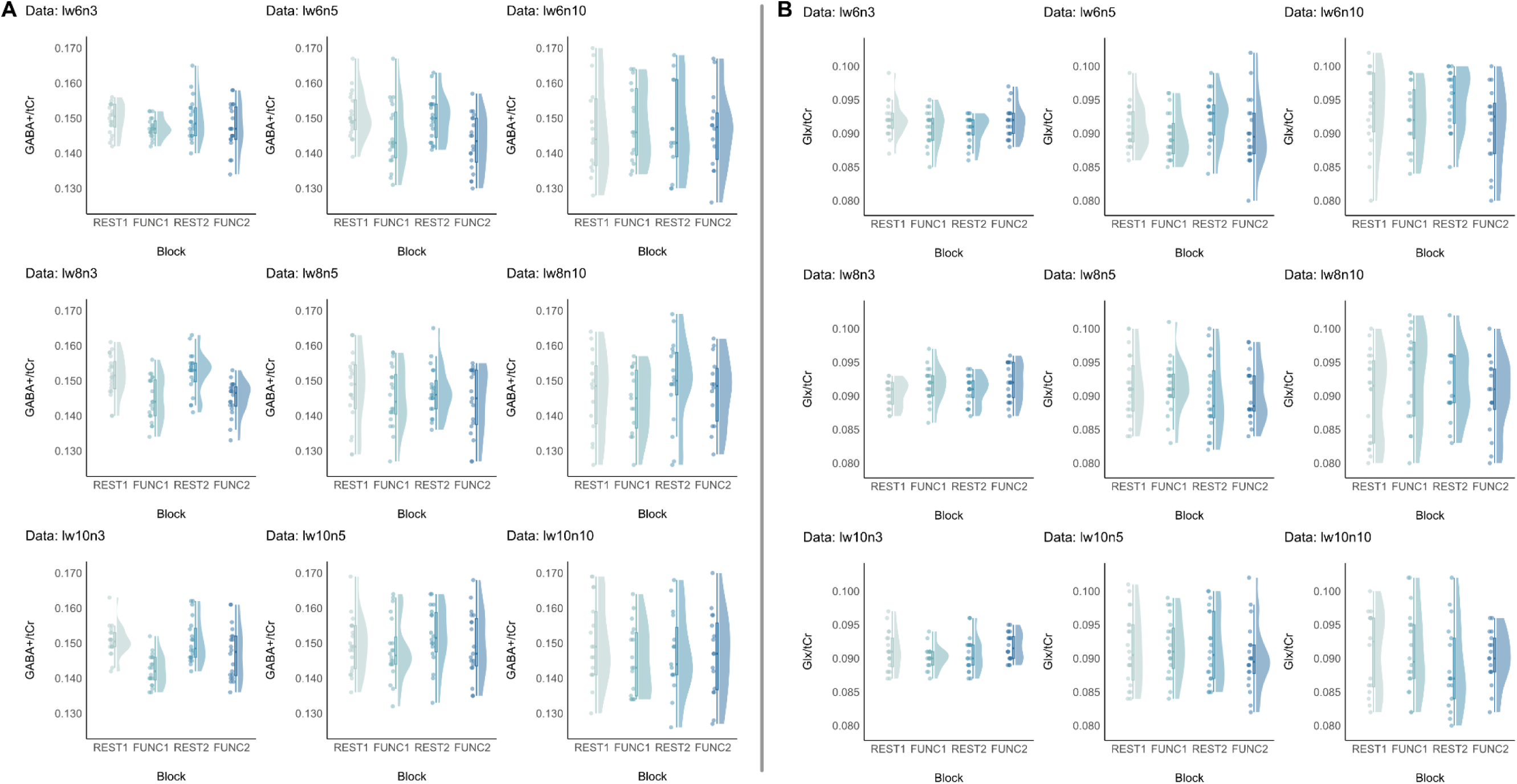
GABA+/tCr and Glx/tCr acquired from block analysis for each data quality level. lw represents linewidth (Hz), and n represents noise_SD_ applied to the simulated dataset. The raincloud plot depicts individual data points, the half-violin plot represents data distribution, and the box plot with whiskers indicates the median and interquartile range. Data that fail quality metrics have not been plotted or included in the analysis.

Overall, ROPE analysis of GABA+/tCr (Figure 10A) and GABA+ (i.u.) (Supplementary Figure 4A) indicated that the reduction of GABA+ during the fMRS blocks was more detectable with good data quality (lw6n3, lw6n5, lw8n3, and lw10n3), as denoted by a lower percentage of the 95% HDI falling inside the ROPE. According to the ROPE analysis, the effect of fMRS blocks on GABA+/tCr and GABA+ (i.u.) can be considered significant in the following cases: a significant effect of both FUNC1 and FUNC2 for the data quality of lw6n5 and lw8n3, and a significant effect of FUNC1 for the data quality of lw10n3, with all effects having high certainty (pd>99%) of being negative and large as expected from the simulated reduction in GABA+ from baseline (see Supplementary Table 5 for more detail).

**Figure 10.**
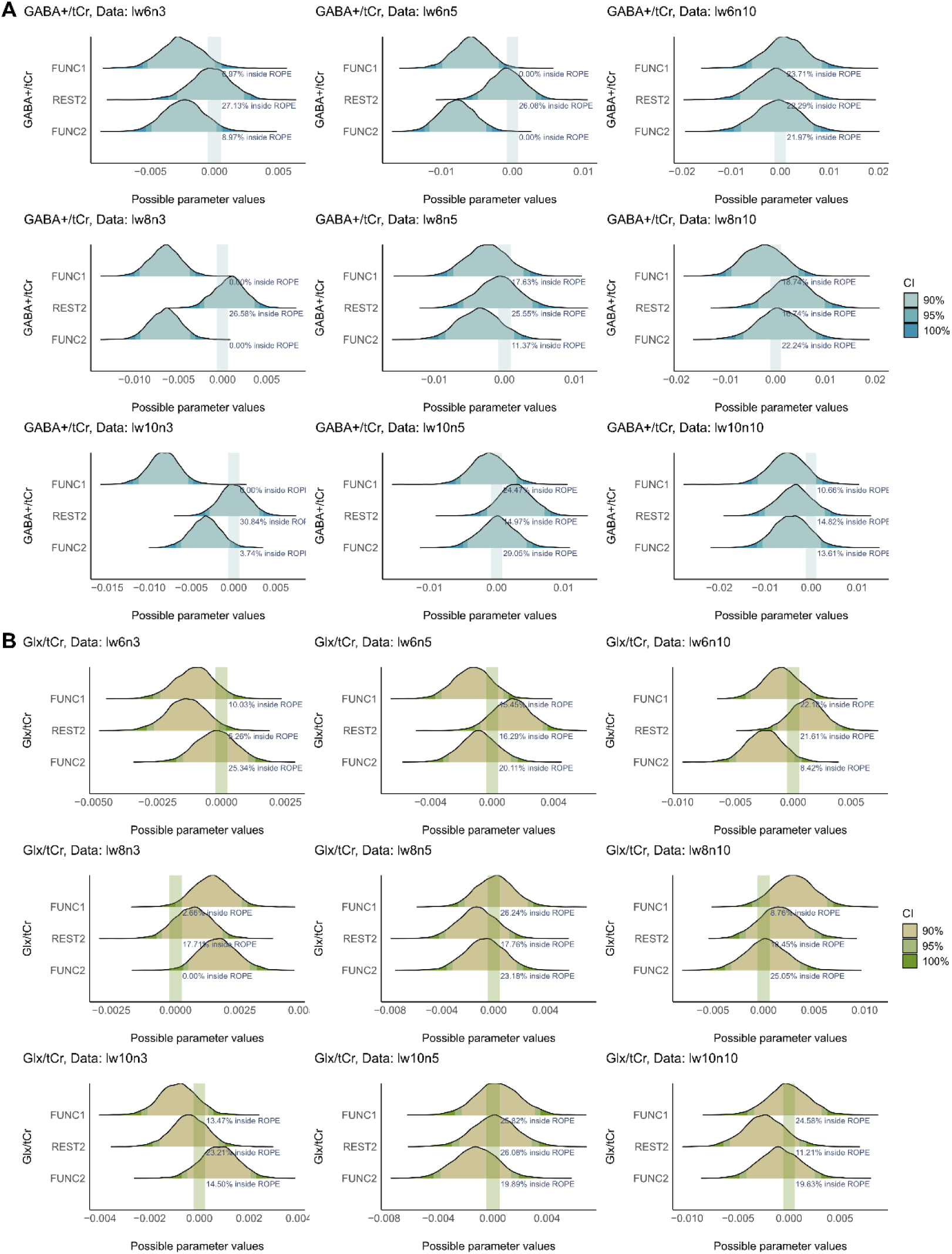
Posterior distribution plots of GABA+/tCr and Glx/tCr of simulated fMRS data analysed with block analysis for varying data quality with different shades representing the range within the posterior distribution that contains a specified proportion (90%, 95% and 100%) of the probability density. The superimposed green boxes show ROPE region (-0.1 to 0.1 times the standard deviation of the outcome variable ^28^ that denotes the range considered practically equivalent to a negligible effect. The percentage displayed shows the portion of the 95% HDI of the posterior distributions that falls within the ROPE.

These ROPE results suggest good agreement between GABA+/tCr and GABA+ (i.u.) quantified with the block analysis method. Interestingly, under the same data quality conditions, the change in GABA+ during FUNC2 is often lower than during FUNC1, as indicated by a higher proportion of the 95% HDI falling inside the ROPE region, suggesting an effect of the simulated phase/frequency drift. While the expected changes in metabolites can be detected at different levels of linewidth, the analyses failed to detect changes in datasets with a high noise_SD_ of 10. This perhaps suggests higher noise levels might have more adverse effects on metabolite quantification than the changes in spectral linewidth, even though both factors were varied within the ranges of realistic MRS data^23,33^.

For Glx/tCr (Figure 10B) and Glx (i.u.) (Supplementary Figure 3A), the expected increase during the fMRS block is less prevalent according to the Bayesian analysis results. Only the effect of FUNC2 on Glx/tCr for the data quality of lw8n3 can be considered as significant (0% of 95%HDI inside ROPE), with a probability of 98.92% of being positive (Median=0.0016, 95% CI [0.0003, 0.003]). This effect is considered to have a large effect size (90.35% probability based on the threshold > 7.18e-04). There was no significant effect of the fMRS blocks on Glx (i.u.) levels for all data quality metrics.

#### Event-related analysis of simulated fMRS data

The quality metrics for each metabolite obtained with the event-related data analysis of simulated fMRS data are presented in Table 3.

**Table 3.** quality metrics of simulated fMRS data included in the event-related analysis for each quality level. lw=line width (Hz), n= noise_SD_. The data presents as mean (SD).

| Data | Metabolite | n | FWHM (Hz) | SNR | Fit Error (%) |
| --- | --- | --- | --- | --- | --- |
| **lw6n3** | Glx | 200 | 8.74 (0.46) | 4.64 (1.51) | 12.16 (2.46) |
|  | GABA | 200 | 17.46 (1.02) | 3.49 (1.16) | 16.18 (3.16) |
|  | NAA | 200 | 4.26 (0.03) | 68.57 (21.18) | 1.75 (0.09) |
| **lw6n5** | Glx | 192 | 8.42 (0.69) | 2.89 (0.88) | 18.11 (4.11) |
|  | GABA | 196 | 17.24 (1.57) | 2.1 (0.69) | 25.23 (6.17) |
|  | NAA | 200 | 4.27 (0.06) | 41.14 (12.76) | 2.05 (0.21) |
| **lw6n10** | Glx | 149 | 8.75 (1.88) | 1.61 (0.45) | 31.36 (7.74) |
|  | GABA | 81 | 16.52 (2.25) | 1.39 (0.29) | 35.53 (6.97) |
|  | NAA | 200 | 4.26 (0.13) | 20.67 (6.24) | 3.07 (0.55) |
| **lw8n3** | Glx | 200 | 8.68 (0.45) | 4.6 (1.43) | 12.04 (2.22) |
|  | GABA | 200 | 17.24 (0.90) | 3.44 (1.13) | 16.17 (3.07) |
|  | NAA | 200 | 4.25 (0.03) | 67.91 (21.35) | 1.77 (0.09) |
| **lw8n5** | Glx | 190 | 8.69 (0.87) | 2.87 (0.87) | 18.24 (4.3) |
|  | GABA | 198 | 17.41 (1.65) | 2.1 (0.69) | 25.03 (6.09) |
|  | NAA | 200 | 4.25 (0.06) | 41.12 (12.84) | 2.04 (0.22) |
| **lw8n10** | Glx | 150 | 8.52 (1.32) | 1.61 (0.47) | 31.73 (8.26) |
|  | GABA | 81 | 16.66 (1.63) | 1.37 (0.32) | 36.78 (8.18) |
|  | NAA | 200 | 4.26 (0.10) | 20.78 (6.38) | 3.03 (0.55) |
| **lw10n3** | Glx | 200 | 8.58 (0.53) | 4.62 (1.46) | 12.23 (2.34) |
|  | GABA | 200 | 17.19 (0.96) | 3.43 (1.16) | 16.63 (3.40) |
|  | NAA | 200 | 4.26 (0.03) | 68.5 (21.34) | 1.75 (0.09) |
| **lw10n5** | Glx | 192 | 8.68 (0.84) | 2.87 (0.89) | 18.35 (4.1) |
|  | GABA | 186 | 17.34 (1.65) | 2.15 (0.71) | 24.84 (6.11) |
|  | NAA | 200 | 4.26 (0.05) | 41.2 (12.65) | 2.05 (0.22) |
| **lw10n10** | Glx | 142 | 8.19 (1.13) | 1.63 (0.44) | 31.04 (8.01) |
|  | GABA | 80 | 16.84 (1.79) | 1.39 (0.30) | 35.6 (7.95) |
|  | NAA | 200 | 4.24 (0.11) | 20.8 (6.33) | 3.03 (0.57) |

Overall, the quantification of GABA+/tCr (Figure 11A) and GABA+ (i.u.) (Supplementary Figure 5A) obtained with event-related analysis revealed an increase in %fit error of GABA+ at higher levels of noise_SD_. This led to the exclusion of data points based on our first criterion (excluding data if %fit error exceeds 50%). Specifically, ON events were predominantly affected by data quality issues in lw6n10, lw8n10, and lw10n10. Visually, GABA+/tCr and GABA+ (i.u.) generally exhibited similar data distributions. Based on the Kruskal-Wallis test, a significant difference in GABA+ levels (p < 0.05) was observed in datasets of lw6n3, lw8n3, lw8n5, lw10n3, and lw10n5. Due to the minimal number of datapoints remaining, the GABA+ datasets with data quality of lw6n10, lw6n10 and lw10n10 were removed from further Bayesian analysis.

**Figure 11.**
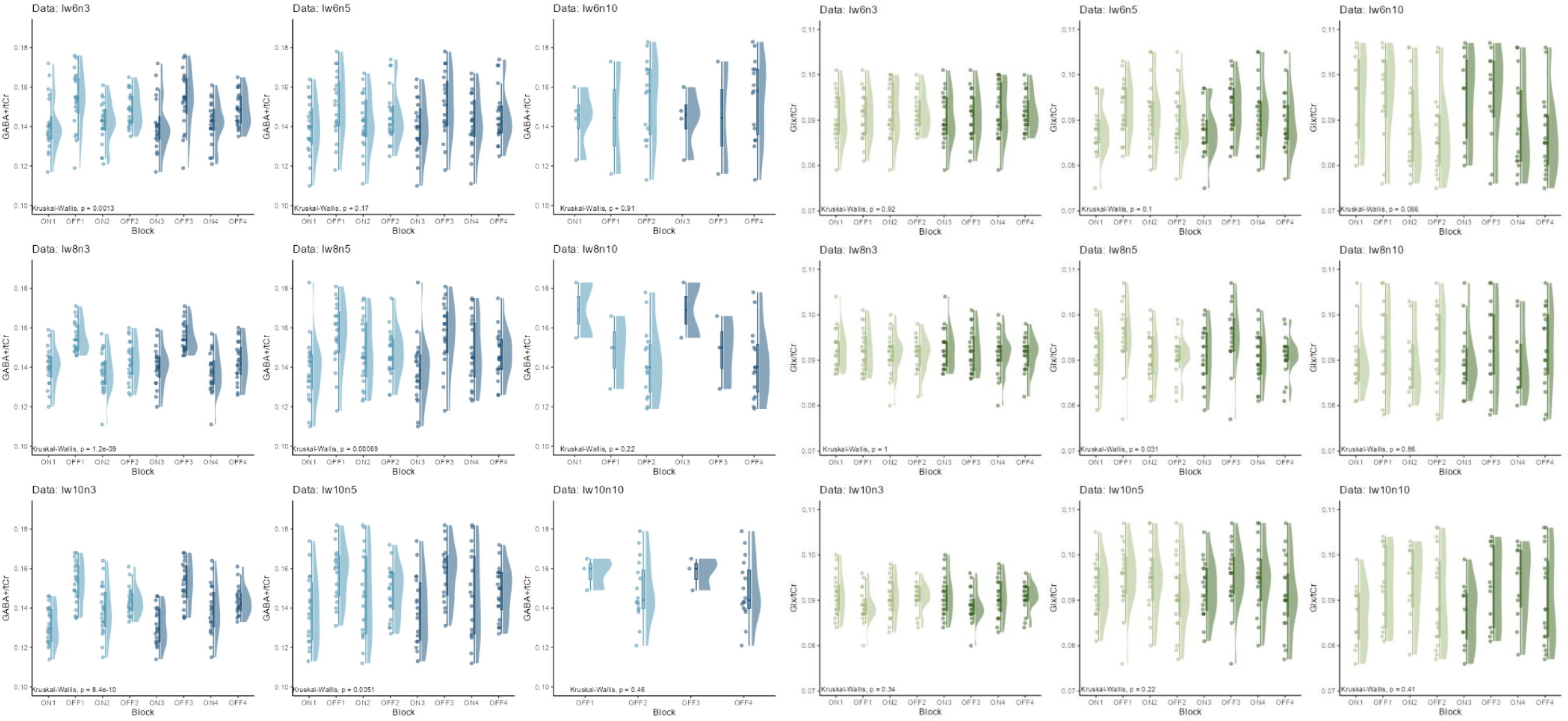
(A) GABA+/tCr and (B) Glx/tCr of simulated fMRS data obtained with event-related analysis based on the stimulus onset of 2s, the data were grouped in half to retain SNR for metabolite quantification. FUNC1 halved into ON1, ON2, OFF1, OFF2; FUNC2 halved into ON3, ON4, OFF3 and OFF. The raincloud plot depicts individual data points, the half-violin plot represents data distribution, and the box plot with whiskers indicates the median and interquartile range. Data that fail quality metrics have not been plotted or included in the analys

Compared to GABA+, both Glx/tCr (Figure 11B) and Glx (i.u.) (Supplementary Figure 5B) obtained with event-related analysis are suggested to be less affected by data quality. This is based on fewer data points being rejected by the quality assurance criteria. However, no significant difference in Glx levels was found across the fMRS blocks when tested with the Kruskal-Wallis test.

Based on the Bayesian analysis, the results generally showed good agreement between GABA+/tCr (Figure 12A) and GABA+ (i.u.) (Supplementary Figure 6A). The results mostly demonstrated the expected reduction in GABA+ compared to baseline during the stimulus-ON TRs. All stimulus-ON blocks (ON1, ON2, ON3, and ON4) could be considered significant for data quality of lw6n3, lw6n5, and lw10n3, with 0% of 95%HDI inside the ROPE region. For data quality of lw8n5, only ON1 and ON3 showed a significant effect.

**Figure 12.**
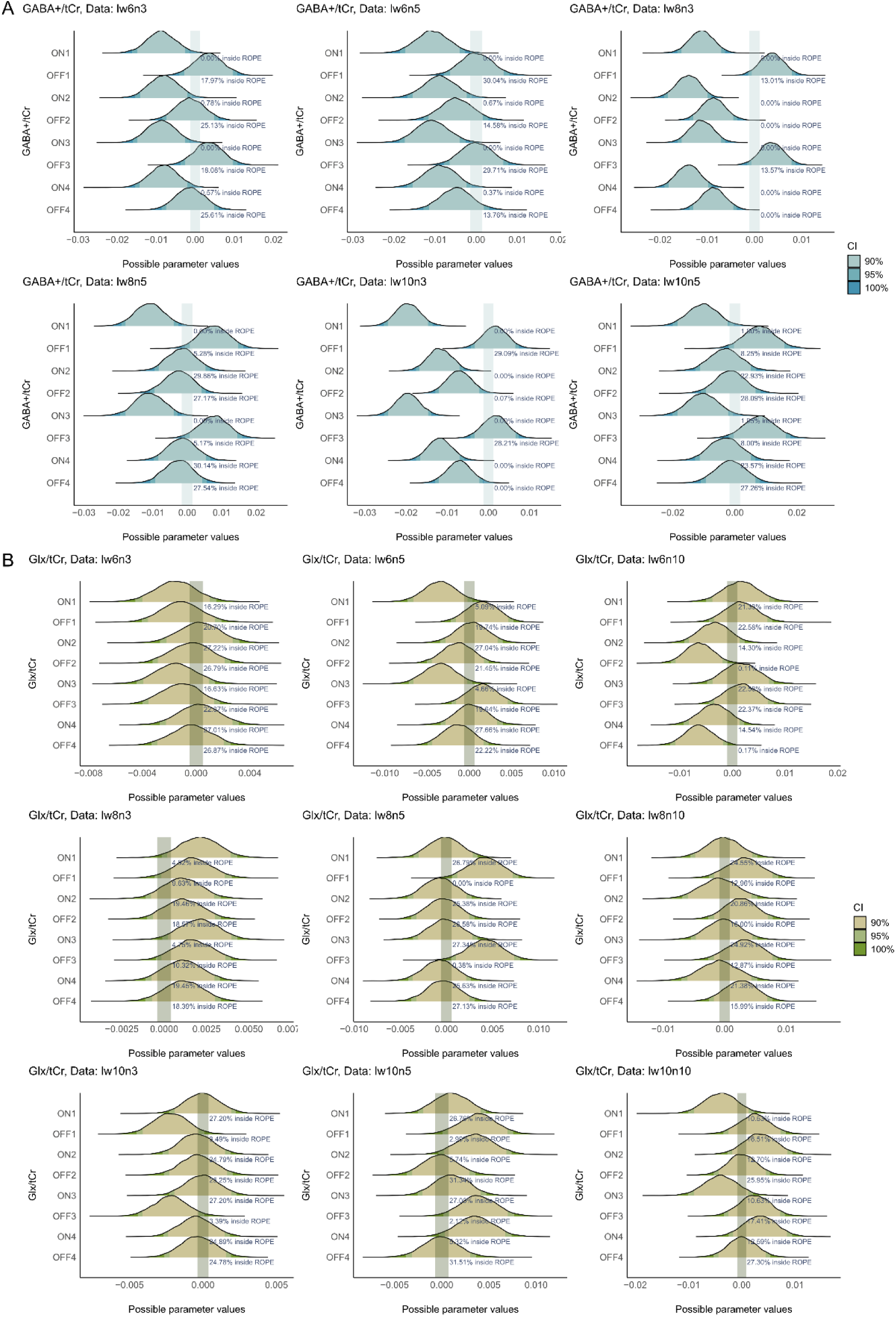
ROPE analysis for (A) GABA+/tCr and (B) Glx/tCr of simulated fMRS data obtained with event-related analysis. FUNC1 halved into ON1, ON2, OFF1, OFF2; FUNC2 halved into ON3, ON4, OFF3 and OFF4.

Surprisingly, datasets with low standard deviation of noise, particularly n3, exhibited a significant effect on both GABA+/tCr and GABA+ (i.u.) during the stimulus-OFFs periods. This was observed for lw8n3, where all parameters, including the TRs during stimulus-OFFs, showed a significant effect on GABA+, except for OFF1 and OFF3. Additionally, results for lw10n3 showed a potentially significant effect of OFF2 on GABA+ (i.u.), with only 1% of the 95% HDI inside the ROPE and significant effect of OFF2 and OFF4 on GABA+/tCr. Here, we hypothesise that at lower noise levels and with fewer transients used in the analysis, linewidth could drive false-positive results, while increasing noise levels lead to overall worsened metabolite quantification.

Glx, on the other hand, mostly showed no significant effect of stimulus-ON (Figure 12B and Supplementary Figure 6B). Surprisingly, there were significant effects of OFF2 and OFF4 on Glx/tCr for data quality of lw6n10 (0.11% and 0.17% inside ROPE, respectively) which also observed in Glx (i.u.). There also significant effects of OFF1 (0% inside ROPE) and OFF3 (0.38% inside ROPE) on Glx/tCr for lw8n5 but this were not observed for Glx (i.u.).

### Sliding window analysis of simulated fMRS data

The quality metrics of data included in the sliding window analysis of simulated data are shown in Table 4. The plots visualised GABA+/tCr and Glx/tCr levels acquired with sliding window analysis approach are shown in Figure 13 and GABA+(i.u.) and Glx (i.u.) are shown in Supplementary Figure 7.

**Figure 13.**
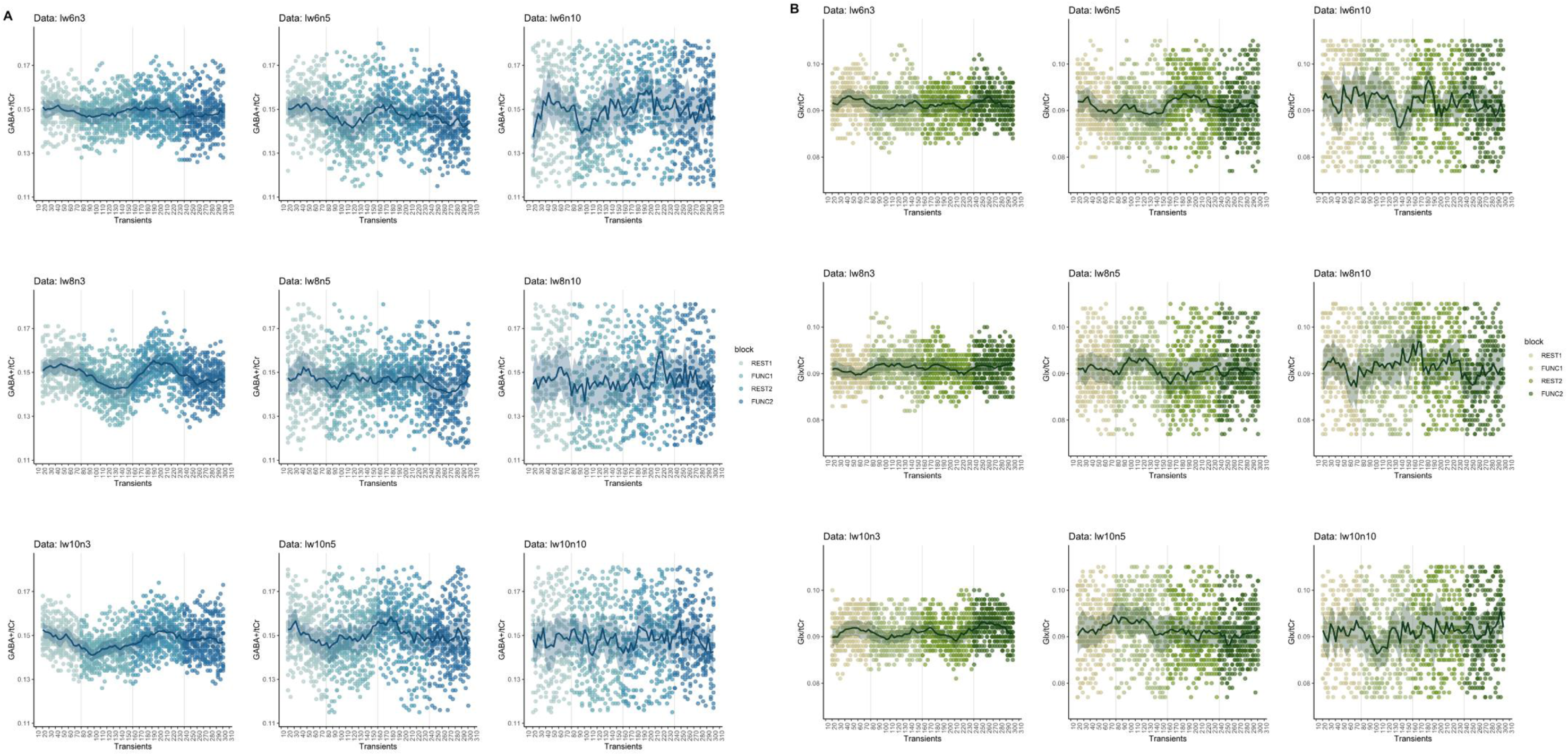
Metabolite levels relative to tCr, acquired using sliding window analysis with varying transient sizes (ww). (A) GABA+/tCr, (B) Glx/tCr. The solid line represents the mean metabolite level, while the shaded area indicates the 95% confidence interval for each window.

**Table 4.** quality metrics of simulated fMRS data included in the sliding window analysis for each quality level. lw=line width (Hz),n= noise_SD_.

| Data | Metabolite | n | FWHM (Hz) | SNR | Fit error (%) |
| --- | --- | --- | --- | --- | --- |
| **lw6n3** | Glx | 1420 | 8.75 (0.39) | 5.10 (0.30) | 10.65 (0.72) |
|  | GABA | 1420 | 17.51 (0.72) | 3.86 (0.20) | 14.07 (0.90) |
|  | NAA | 1420 | 4.26 (0.03) | 75.71 (3.25) | 1.71 (0.06) |
| **lw6n5** | Glx | 1412 | 8.61 (0.69) | 3.11 (0.25) | 16 (1.51) |
|  | GABA | 1401 | 17.42 (1.17) | 2.30 (0.15) | 21.59 (1.65) |
|  | NAA | 1420 | 4.26 (0.05) | 45.22 (2.01) | 1.92 (0.10) |
| **lw6n10** | Glx | 1149 | 8.62 (1.23) | 1.66 (0.22) | 29.03 (3.99) |
|  | GABA | 1056 | 17.16 (2.03) | 1.19 (0.12) | 40.17 (4.24) |
|  | NAA | 1420 | 4.27 (0.09) | 22.81 (0.89) | 2.71 (0.16) |
| **lw8n3** | Glx | 1420 | 8.69 (0.36) | 5.12 (0.29) | 10.64 (0.73) |
|  | GABA | 1420 | 17.50 (0.78) | 3.85 (0.20) | 14.14 (0.93) |
|  | NAA | 1420 | 4.26 (0.03) | 75.12 (3.03) | 1.71 (0.06) |
| **lw8n5** | Glx | 1385 | 8.62 (0.71) | 3.10 (0.22) | 16.00 (1.41) |
|  | GABA | 1386 | 17.25 (1.22) | 2.32 (0.16) | 21.41 (1.65) |
|  | NAA | 1420 | 4.26 (0.04) | 45.34 (2.03) | 1.92 (0.09) |
| **lw8n10** | Glx | 1126 | 8.58 (1.12) | 1.65 (0.22) | 28.99 (3.9) |
|  | GABA | 993 | 17 (1.91) | 1.17 (0.12) | 40.49 (4.21) |
|  | NAA | 1420 | 4.26 (0.09) | 22.93 (0.99) | 2.68 (0.16) |
| **lw10n3** | Glx | 1420 | 8.67 (0.42) | 5.10 (0.30) | 10.70 (0.68) |
|  | GABA | 1420 | 17.39 (0.76) | 3.84 (0.20) | 14.21 (0.83) |
|  | NAA | 1420 | 4.26 (0.03) | 75.4 (3.38) | 1.71 (0.07) |
| **lw10n5** | Glx | 1371 | 8.63 (0.67) | 3.11 (0.24) | 15.95 (1.52) |
|  | GABA | 1381 | 17.50 (1.23) | 2.33 (0.17) | 21.26 (1.73) |
|  | NAA | 1420 | 4.26 (0.05) | 45.34 (2.05) | 1.92 (0.09) |
| **lw10n10** | Glx | 1061 | 8.69 (1.21) | 1.64 (0.21) | 29.14 (4.16) |
|  | GABA | 1123 | 16.98 (1.78) | 1.19 (0.12) | 39.83 (3.86) |
|  | NAA | 1420 | 4.25 (0.09) | 22.87 (0.95) | 2.67 (0.17) |

Visually, GABA+/tCr and GABA+ (i.u.) determined by sliding window analysis showed a trend of the expected decrease in GABA+ during the fMRS blocks, which is particularly evident for the data quality of lw8n3. The same trend was also observed in GABA+ (i.u.). For Glx, a pattern reflecting an increase in Glx/tCr and Glx (i.u.) levels was also observed, albeit only subtly.

One notable feature from the plot was that as the standard deviation of the data increased, the data distributions for both GABA+ and Glx became almost uniform across all time points, reflecting that increased noise interfered with the reliability of metabolite quantification. On the other hand, at the same level of noise, the linewidth showed less effect on metabolite quantification.

ROPE results from Bayesian analyses are shown for GABA+/tCr in Figure 14*A*, GABA+ (i.u.) in Supplementary Figure 8A, Glx/tCr Figure 14B in and Glx (i.u.) in Supplementary Figure 8B. The data showed high multicollinearity between parameters, particularly for FUNC2 and REST2 across several data qualities. The multicollinearity between parameters invalidates the ROPE due to the parameters moving towards or away from the ROPE and altering the proportion distribution^34^. Multicollinearity with correlations greater than 0.7 between REST2 and FUNC2 were observed in the following datasets: lw6n3 (r = 0.71), lw8n3 (r = 0.73), lw8n5 (r = 0.77), lw10n2 (r = 0.75), and lw10n5 (r = 0.73). We hypothesise that this was due to the simulated frequency drift and negative phase drift, which might have had a more pronounced effect on transients towards the end of the simulated spectra.

**Figure 14.**
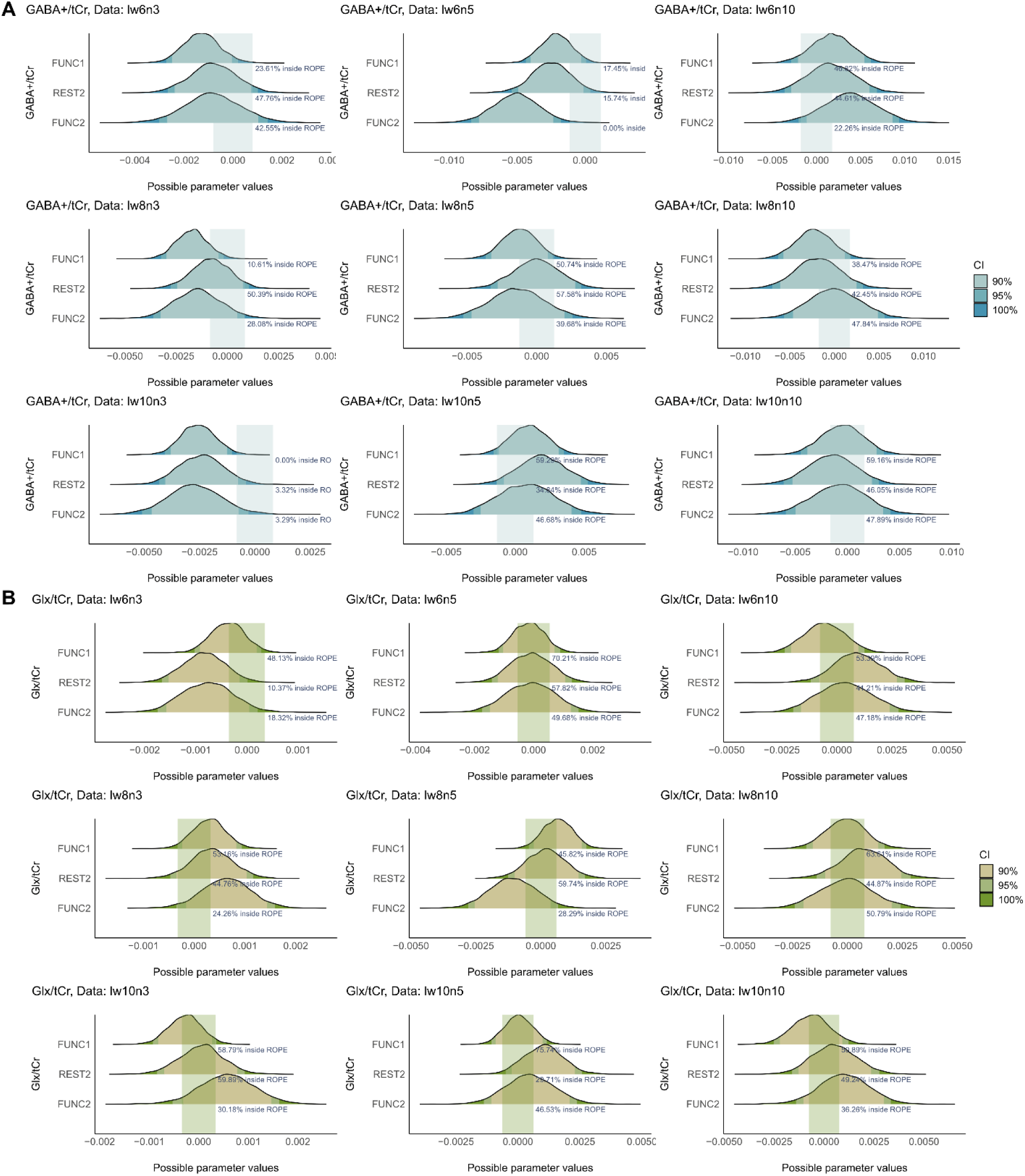
ROPE analysis for (A) GABA+/tCr and (B) Glx/tCr ratios obtained using sliding window analysis. Multicollinearity (r> 0.7) between REST2 and FUNC2 is observed in some of datasets. For GABA+/tCr, multicollinearity is observed in lw6n3 (r = 0.71), lw8n3 (r = 0.73), lw8n5 (r = 0.77), lw10n3 (r = 0.75), and lw10n5 (r = 0.73); for Glx/tCr, multicollinearity is observed in lw6n3 (r = 0.74), lw8n3 (r = 0.74), lw8n5 (r = 0.76), lw10n3 (r = 0.77), and lw10n5 (r = 0.72).

## Discussion

In this study, we investigated the effect of the number of transients on metabolite quantification and data quality on fMRS quantification of GABA+ and glutamate. Here, we utilised the publicly available Big GABA MEGA-PRESS dataset to explore the effects of the choice of the number of transients and analysis methods. We then examined the effects of data quality on metabolite quantification using a simulated MEGA-PRESS dataset with known ground truth changes in metabolite levels.

### Analysis of baseline data

Using the Big GABA dataset as baseline, the results suggested that the choice of the number of transients and analysis method could lead to a false positive increase in measured GABA+ levels, even in the absence of a stimulus, while Glx is less affected.

When using the metabolite quantified using all transients as baseline, GABA+/tCr and GABA+(i.u.) exhibited only moderate correlation (r ≥ 0.6) from 64 transients and higher, whereas Glx/tCr and Glx (i.u.) showed a strong correlation (r ≥ 0.7) with reference values starting from a binning size of 32 transients. These results suggest that shorter scan times could suffice for Glx.

One important finding was observed in sliding window analysis, where a window width of 16 transients significantly increased both GABA+/tCr and GABA+ (i.u.) despite data having been acquired at baseline. Here we hypothesise that GABA+/tCr and GABA+ (i.u.) obtained with the sliding window analysis potentially incorporate the temporal variability from the previous timepoint into the quantification, which could propagate through the analysis, leading to a false positive changes in GABA+. One potential explanation for the error being more in the positive direction could be that the fitting algorithm, which uses a simple Gaussian to fit, with a linear baseline, incorporates noise from regions of the spectrum surrounding the GABA+ peak and potentially includes the effect of phase/frequency drift that causes a shift in the baseline. Overall, results align with theoretical relationships indicating that the model fit error has an inverse square root relationship with the number of transients and is inversely proportional to SNR ^31^.

Despite anticipating stable metabolite levels, variations in GABA+ and Glx without stimulation cannot be ruled out, as indicated by a study that found metabolic changes in the visual cortex under eyes-closed conditions ^35^. Interestingly, the same study investigated the Big GABA dataset using a sliding window analysis and found no change in GABA+ and glutamate using a window width of 128 with a step size of 2 transients, which aligns with our results of no significant change in GABA+ at the same window width. This could be due to the high number of transients and thus sufficient SNR for reliable quantification.

These results highlight critical implications in balancing between scan time/temporal resolution and quantification reliability, particularly for GABA measurement. Careful consideration is especially critical for GABA+ quantification due to additional potential SNR loss in GABA-edited MRS compared to non-editing techniques e.g. PRESS. This loss can arise from imperfect subtraction of the edit-on and edit-off spectra, which might be further affected by frequency and phase instabilities, MM-contamination, scanner instabilities and motion ^36,37^. These insights apply not only to high-temporal resolution analysis but also to static MRS experimental design, where selecting an adequate number of transients is crucial for minimising scan time while ensuring metabolite quantification accuracy.

### Analysis of simulated fMRS data

Here we also investigated the effects of various analysis methods on simulated fMRS data. The dataset incorporated known changes in GABA+ and Glx levels, altered by three different linewidths and three different noise_SD_ levels to simulate real-world conditions. We hypothesised a 20% decrease in GABA+ and a 20% increase in Glx during fMRS blocks in response to tactile stimulation, reflecting findings from ^17^. This relatively large effect size was chosen to thoroughly examine the role of analysis methods. The range of linewidths was chosen to mimic a typical spectrum that has a linewidth of 5–10 Hz, and levels of noise_SD_ were chosen to reflect scenarios of data with low, medium and high noise scenarios ^23,33^.

Our investigation revealed that noise has a more pronounced impact on metabolite quantification than linewidth. Specifically, a noise_SD_ level of 10 resulted in a significant amount of data being discarded for GABA+/tCr and GABA+ (i.u.) due to fitting errors exceeding 50%. We hypothesise that the high fitting errors for GABA+ are due to the inherently small and complicated line shape of GABA+ peaks. Additionally, the GABA-edited difference spectrum has been demonstrated to have potential GABA+ signal loss of up to 40% due to incomplete subtraction between the edit-ON and edit-OFF spectra from various sources such as frequency and phase instability, MM- contamination, motion and scanner instabilities (Evans et al., 2013; Harris et al., 2014; Kaiser et al., 2008). In contrast, Glx signals are generally larger and more well-defined due to the higher Glx concentration which makes its quantification more robust and less prone to quantification error.

The type of analysis and the manner in which transients are indexed significantly influence metabolite quantification. Our block analysis captured the expected changes in GABA+ under conditions of low noise (noise_SD_ < 3) and typical linewidth (6-8 Hz). These results also showed that block analysis could capture the averaged changes of these neurometabolites within each fMRS block, despite the interleaving of metabolite changes during the fMRS blocks.

Event-related analysis, which indexes transients in a stimulus-locked manner, robustly detected reductions in GABA+ during all stimulus-ON phases across datasets with good data quality of noise < 10. The event-related results indicate a sensitivity to the low number of transients used for the analysis. Interestingly, we also found GABA+ changes during the stimulus-OFFs for datasets with relatively low noise but broader linewidth, particularly n3lw8 and n3lw10. Notably, Glx showed no significant changes across all stimulus-ONs, although certain high-noise simulations (lw8n5 and lw6n10) showed a potential false positive change during stimulus-OFFs. These results suggest that linewidths influence the accuracy of the GABA+ measurement while high noise levels adversely affect Glx quantification, leading to spurious changes being detected during stimulus-OFF periods. One potential explanation is that Glx is more affected by line broadening due to reduced separation of its double peaks, whereas the broader linewidth of GABA+ may introduce more noise into the simple Gaussian model used for fitting, increasing the likelihood of fitting errors. However, further studies are needed to test this hypothesis.

Our sliding window analysis introduced additional complexities. High multicollinearity (r > 0.7) between parameters towards the experiment’s end suggested that the nature of sliding window analysis—overlaying data points—might influence how the results are interpretated. Overall, the effect sizes of all blocks analysed with sliding windows are small. While the decrease in GABA+ is consistent with the expected changes from simulated GABA+ levels, the directions of Glx changes are mixed. It should also be noted that we applied our change in GABA in a stimulus-locked manner whereas in reality, the metabolite response function is likely delayed, with its timing yet unknown (also see ^7^).

Our reproducibility results align with previous studies demonstrating that test-retest coefficients of variance for glutamate begin to stabilize at 8–16 transients for short TE semi-LASER^38^. Similarly, another study reported minimal improvement in the %fit error of GABA in the sensorimotor cortex when increasing the number of transients beyond 40^31^. Consistent with these findings, our results suggest the potential for achieving higher temporal resolution in fMRS compared to the conventionally used range of 64–256 transients. However, this is contingent on data quality, with lower noise levels and a spectral linewidth of approximately 6–8 Hz being optimal. This is in agreement with the expert consensus, suggesting that a linewidth exceeding 0.1 ppm (12.77 Hz for 3 T) is considered low-quality data (Wilson et al., 2019).

Overall, our results suggest that while it is possible to reduce the number of transients used for Glx for both block and event-related analysis, the choice of the number of transients has a more significant effect on GABA+. In our current study, the smallest number of transients chosen for measuring GABA+ with a spectral-editing sequence without any substantial effect on its quantification was 32, based on high Pearson correlation values observed in our baseline data analyses. Results also indicate that data quality is crucial, as we would expect. Lower noise and a linewidth of approximately 6-8 Hz are preferable, especially for analysis methods utilising fewer transients.

Our results indicate that different analytical approaches—block, event-related, and sliding window analysis—capture unique dynamics in metabolite responses. Specifically, block analysis has shown that changes in metabolites with a temporal resolution of 2 seconds can be adequately detected with this method. Additionally, employing transients time locked to stimulus presentation or task in event-related analysis enhances the temporal resolution and specificity of metabolite responses to stimuli. Sliding window analysis, often used as an exploratory method, offers the highest effective temporal resolution. However, caution is necessary when analysing and interpreting data using this method, particularly due to the potential for collinearity among data points caused by overlap.

### Limitations of the current study

This study has some limitations. The first limitation is that these results may be specific to spectral editing approaches, particularly MEGA-PRESS, as used in the current study. Given that spectral editing techniques can lead to a reduction in SNR due to spectral subtraction, other fMRS studies employing non-spectral editing methods might achieve reliable results with even fewer transients than demonstrated here.

In terms of our simulated data, while the FID-A simulation for MEGA-PRESS spectra allowed for the simulation of ’clean’ data with controlled parameters and spectral imperfections, it does not perfectly mirror real-world conditions. The simulated MM-peaks, modelled using simple Gaussian peaks, may not accurately represent the complex nature of MM peaks found in MRS.

Our analysis of simulated data revealed the absence of Glx changes across different analysis approaches, despite being present in the simulated fMRS block. This discrepancy may stem from the possibility that the Glx peaks simulated in this study are too narrow compared to real-world *in vivo* Glx data. Real-world data typically exhibit slightly broader peaks that include multiple underlying MM-peaks^39,40^. The use of lower line broadening and simulated MM peaks could potentially lead to the underestimation of Glx and explain the lower quantified Glx values compared to GABA+ seen in our dataset.

It is also conceivable that the results observed here may be specific to the fitting algorithm used. While there has been no investigation into the comparison of the effect of model choice on Glx peak between using a singlet Gaussian, which ignores peak splitting, or using doublet Gaussian peaks as implemented in Gannet, both should roughly agree in trend changes. Although beyond the scope of this study, exploring the effects of different MRS fitting algorithms on various analysis approaches would be of interest, yet no gold standard for fMRS analysis exists at this time.

## Conclusion

As with all fMRS or MRS studies, careful consideration in experimental design and analysis is paramount. The right balance in the trade-off between data quality and optimal temporal resolution is essential for achieving meaningful results, to answer each specific research question. We advocate for the adoption of a reporting consensus for (f)MRS studies such as MRS-Q and MRSinMRS^41,42^ and conservative approaches to the number of transients used in metabolite quantification, especially for GABA. Although a specific reporting standard for fMRS has not yet been established, adopting these minimal reporting standards can significantly enhance scientific integrity, advancing the field through ensuring reliable and reproducible research.

## Data Availability

All data produced in the present study are available upon reasonable request to the authors

https://osf.io/pfdwj/

## Acknowledgements

NP is supported by a Simons SFARI Human Cognitive and Behavioral Science award. NP is supported through the MRC Centre for Neurodevelopmental Disorders. This study represents independent research in part funded by the National Institute for Health and Care Research (NIHR) Maudsley Biomedical Research Centre (BRC) at South London and Maudsley NHS Foundation Trust and King’s College London. DP is supported by Chaing Mai University.

## Abbreviations

lw: line width
n: noise
Glx: Glutamine and Glutamate
GABA: Gamma-aminobutyric Acid
GABA+: Gamma-aminobutyric acid and macromolecules
SNR: Signal to noise ratio

## References

1. Mullins PG. Towards a theory of functional magnetic resonance spectroscopy (fMRS): A meta-analysis and discussion of using MRS to measure changes in neurotransmitters in real time. Scand J Psychol. 2018;59(1):91–103. doi:10.1111/sjop.12411

2. Koolschijn RS, Clarke WT, Ip IB, Emir UE, Barron HC. Event-related functional magnetic resonance spectroscopy. NeuroImage. 2023;276:120194. doi:10.1016/j.neuroimage.2023.120194

3. Pasanta D, He JL, Ford T, Oeltzschner G, Lythgoe DJ, Puts NA. Functional MRS studies of GABA and glutamate/Glx – A systematic review and meta-analysis. Neurosci Biobehav Rev. 2023;144:104940. doi:10.1016/j.neubiorev.2022.104940

4. Lin A, Andronesi O, Bogner W, et al. Minimum Reporting Standards for in vivo Magnetic Resonance Spectroscopy (MRSinMRS): Experts’ consensus recommendations. NMR Biomed. 2021;34(5):e4484. doi:10.1002/nbm.4484

5. Peek AL, Rebbeck TJ, Leaver AM, et al. A comprehensive guide to MEGA-PRESS for GABA measurement. Anal Biochem. 2023;669:115113. doi:10.1016/j.ab.2023.115113

6. Mikkelsen M, Loo RS, Puts NAJ, Edden RAE, Harris AD. Designing GABA-Edited Magnetic Resonance Spectroscopy Studies: Considerations of Scan Duration, Signal-To-Noise Ratio and Sample Size. J Neurosci Methods. 2018;303:86–94. doi:10.1016/j.jneumeth.2018.02.012

7. Pasanta D, Powell H, Hafeez N, Lythgoe DJ, Puts NA. Decoupling of GABA and Glutamine-Glutamate Dynamics and their role in tactile perception: An fMRS Study. Published online November 28, 2024:2024.11.28.625809. doi:10.1101/2024.11.28.625809

8. Mikkelsen M, Rimbault DL, Barker PB, et al. Big GABA II: Water-referenced edited MR spectroscopy at 25 research sites. NeuroImage. 2019;191:537–548. doi:10.1016/j.neuroimage.2019.02.059

9. Simpson R, Devenyi GA, Jezzard P, Hennessy TJ, Near J. Advanced processing and simulation of MRS data using the FID appliance (FID-A)-An open source, MATLAB-based toolkit. Magn Reson Med. 2017;77(1):23–33. doi:10.1002/mrm.26091

10. Zhang Y, An L, Shen J. Fast computation of full density matrix of multispin systems for spatially localized in vivo magnetic resonance spectroscopy. Med Phys. 2017;44(8):4169–4178. doi:10.1002/mp.12375

11. Govindaraju V, Young K, Maudsley AA. Proton NMR chemical shifts and coupling constants for brain metabolites. NMR Biomed. 2000;13(3):129–153. doi:10.1002/1099-1492(200005)13:3<129::AID-NBM619>3.0.CO;2-V

12. Lymer K, Haga K, Marshall I, Sailasuta N, Wardlaw J. Reproducibility of GABA measurements using 2D *J*-resolved magnetic resonance spectroscopy. Magn Reson Imaging. 2007;25(5):634–640. doi:10.1016/j.mri.2006.10.010

13. Near J, Andersson J, Maron E, et al. Unedited in vivo detection and quantification of γ-aminobutyric acid in the occipital cortex using short-TE MRS at 3 T. NMR Biomed. 2013;26(11):1353–1362. doi:10.1002/nbm.2960

14. Oeltzschner G, Zöllner HJ, Hui SCN, et al. Osprey: Open-source processing, reconstruction & estimation of magnetic resonance spectroscopy data. J Neurosci Methods. 2020;343:108827. doi:10.1016/j.jneumeth.2020.108827

15. Craven AR, Bhattacharyya PK, Clarke WT, et al. Comparison of seven modelling algorithms for γ-aminobutyric acid–edited proton magnetic resonance spectroscopy. NMR Biomed. 2022;35(7). doi:10.1002/nbm.4702

16. Stanley JA, Raz N. Functional Magnetic Resonance Spectroscopy: The “New” MRS for Cognitive Neuroscience and Psychiatry Research. Front Psychiatry. 2018;9:76. doi:10.3389/fpsyt.2018.00076

17. Heba S, Puts NAJ, Kalisch T, et al. Local GABA Concentration Predicts Perceptual Improvements After Repetitive Sensory Stimulation in Humans. Cereb Cortex. 2016;26(3):1295–1301. doi:10.1093/cercor/bhv296

18. Hui SCN, Mikkelsen M, Zöllner HJ, et al. Frequency drift in MR spectroscopy at 3T. NeuroImage. 2021;241:118430. doi:10.1016/j.neuroimage.2021.118430

19. Matlab S. Matlab. MathWorks Natick MA. 2012;9.

20. Edden RAE, Puts NAJ, Harris AD, Barker PB, Evans CJ. Gannet: A batch-processing tool for the quantitative analysis of gamma-aminobutyric acid–edited MR spectroscopy spectra. J Magn Reson Imaging. 2014;40(6):1445–1452. doi:10.1002/jmri.24478

21. Ernst T, Chang L. Elimination of artifacts in short echo time 1H MR spectroscopy of the frontal lobe. Magn Reson Med. 1996;36(3):462–468.

22. Wilson M, Andronesi O, Barker PB, et al. Methodological consensus on clinical proton MRS of the brain: Review and recommendations. Magn Reson Med. 2019;82(2):527–550. doi:10.1002/mrm.27742

23. Mikkelsen M, Barker PB, Bhattacharyya PK, et al. Big GABA: Edited MR spectroscopy at 24 research sites. NeuroImage. 2017;159:32–45. doi:10.1016/j.neuroimage.2017.07.021

24. Kruschke JK. Bayesian Analysis Reporting Guidelines. Nat Hum Behav. 2021;5(10):1282–1291. doi:10.1038/s41562-021-01177-7

25. Bürkner PC. brms: An R Package for Bayesian Multilevel Models Using Stan. J Stat Softw. 2017;80:1-28. doi:10.18637/jss.v080.i01

26. Makowski D, Ben-Shachar MS, Lüdecke D. bayestestR: Describing effects and their uncertainty, existence and significance within the Bayesian framework. J Open Source Softw. 2019;4(40):1541.

27. Cohen J. Statistical Power Analysis. Curr Dir Psychol Sci. 1992;1(3):98–101.

28. Kruschke JK. Rejecting or Accepting Parameter Values in Bayesian Estimation. Adv Methods Pract Psychol Sci. 2018;1(2):270–280. doi:10.1177/2515245918771304

29. Brooks WM, Friedman SD, Stidley CA. Reproducibility of 1H-MRS in vivo. Magn Reson Med Off J Int Soc Magn Reson Med. 1999;41(1):193–197.

30. Kreis R. Issues of spectral quality in clinical 1H-magnetic resonance spectroscopy and a gallery of artifacts. NMR Biomed. 2004;17(6):361–381. doi:10.1002/nbm.891

31. Mikkelsen M, Loo RS, Puts NAJ, Edden RAE, Harris AD. Designing GABA-Edited Magnetic Resonance Spectroscopy Studies: Considerations of Scan Duration, Signal-To-Noise Ratio and Sample Size. J Neurosci Methods. 2018;303:86–94. doi:10.1016/j.jneumeth.2018.02.012

32. Terpstra M, Cheong I, Lyu T, et al. Test-retest reproducibility of neurochemical profiles with short-echo, single-voxel MR spectroscopy at 3T and 7T. Magn Reson Med. 2016;76(4):1083–1091. doi:10.1002/mrm.26022

33. Tal A. The future is 2D: spectral-temporal fitting of dynamic MRS data provides exponential gains in precision over conventional approaches. Magn Reson Med. 2023;89(2):499–507. doi:10.1002/mrm.29456

34. Kruschke J. Doing Bayesian data analysis: A tutorial with R, JAGS, and Stan. Published online 2014.

35. Rideaux R. Temporal Dynamics of GABA and Glx in the Visual Cortex. eNeuro. 2020;7(4). doi:10.1523/ENEURO.0082-20.2020

36. Evans CJ, Puts NAJ, Robson SE, et al. Subtraction artifacts and frequency (Mis-)alignment in J-difference GABA editing. J Magn Reson Imaging. 2013;38(4):970–975. doi:10.1002/jmri.23923

37. Rideaux R, Mikkelsen M, Edden RAE. Comparison of methods for spectral alignment and signal modelling of GABA-edited MR spectroscopy data. NeuroImage. 2021;232:117900. doi:10.1016/j.neuroimage.2021.117900

38. Terpstra M, Cheong I, Lyu T, et al. Test-retest reproducibility of neurochemical profiles with short-echo, single-voxel MR spectroscopy at 3T and 7T. Magn Reson Med. 2016;76(4):1083–1091. doi:10.1002/mrm.26022

39. Birch R, Peet AC, Dehghani H, Wilson M. Influence of macromolecule baseline on 1H MR spectroscopic imaging reproducibility. Magn Reson Med. 2017;77(1):34–43. doi:10.1002/mrm.26103

40. Mullins PG, McGonigle DJ, O’Gorman RL, et al. Current practice in the use of MEGA-PRESS spectroscopy for the detection of GABA. NeuroImage. 2014;86:43–52. doi:10.1016/j.neuroimage.2012.12.004

41. Lin A, Andronesi O, Bogner W, et al. Minimum Reporting Standards for in vivo Magnetic Resonance Spectroscopy (MRSinMRS): Experts’ consensus recommendations. NMR Biomed. 2021;34(5):e4484. doi:10.1002/nbm.4484

42. Peek AL, Rebbeck T, Puts NA, Watson J, Aguila MER, Leaver AM. Brain GABA and glutamate levels across pain conditions: A systematic literature review and meta-analysis of 1H-MRS studies using the MRS-Q quality assessment tool. NeuroImage. 2020;210:116532. doi:10.1016/j.neuroimage.2020.116532

