## Supplementary for "Evaluating parameter selection and analysis approaches on quality and reproducibility of functional MRS"

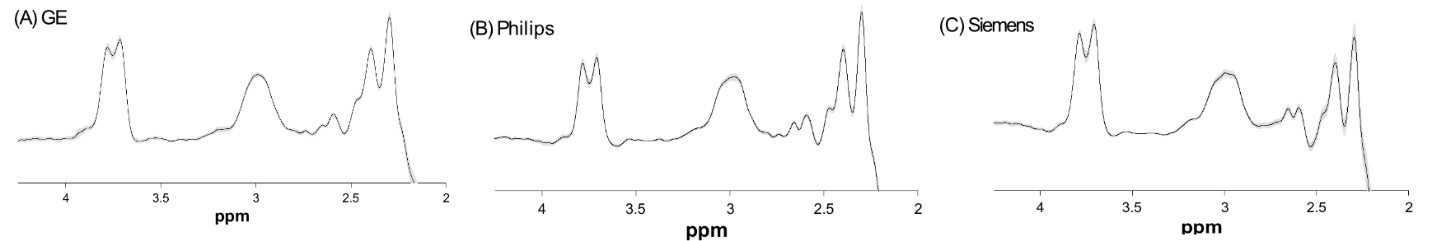

Supplementary Figure 1 Averaged spectra with 95% CI shaded in grey from (A) GE sites (G5 and G7, n=20), (B) Philips sites (P1 and P3), n=20), and (C) Siemens sites (S1 and S3, n=20).

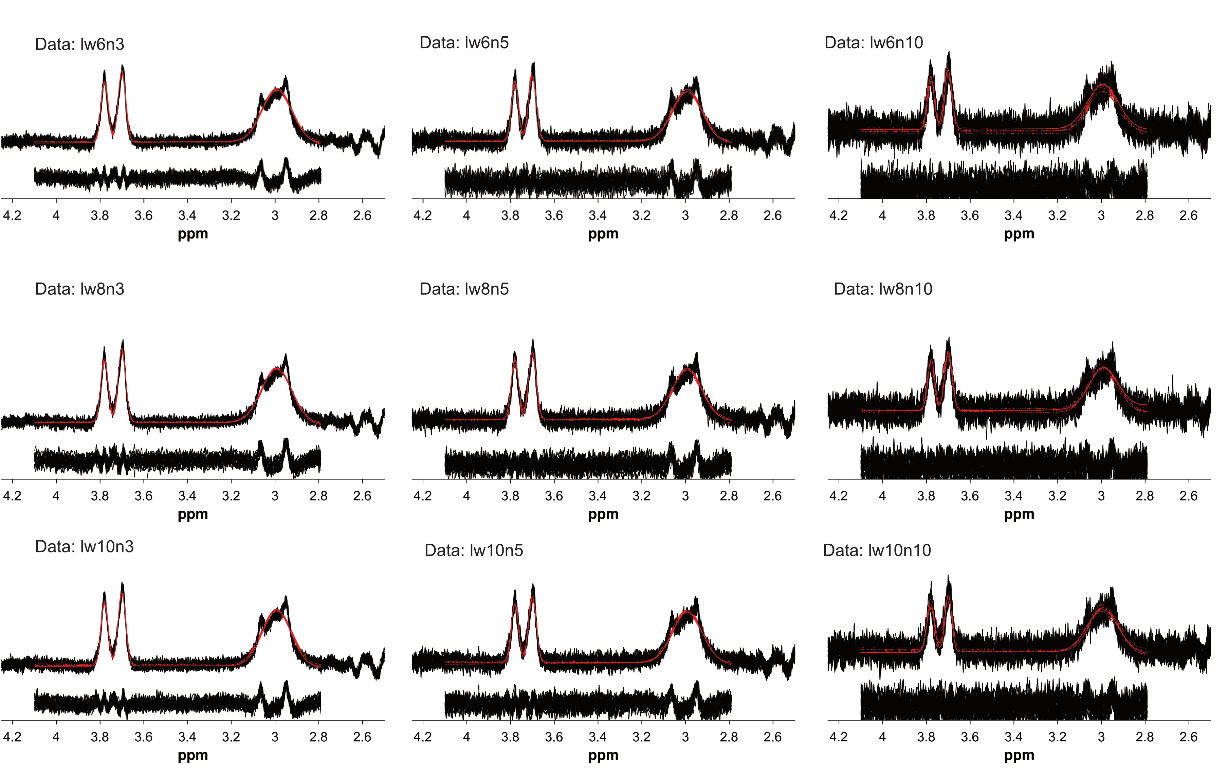

Supplementary Figure 2 All simulated MEGA-PRESS spectra with signal model fitted in red and residual from model fit below the spectra. lw=linewidth(Hz);n=noise_SD_.

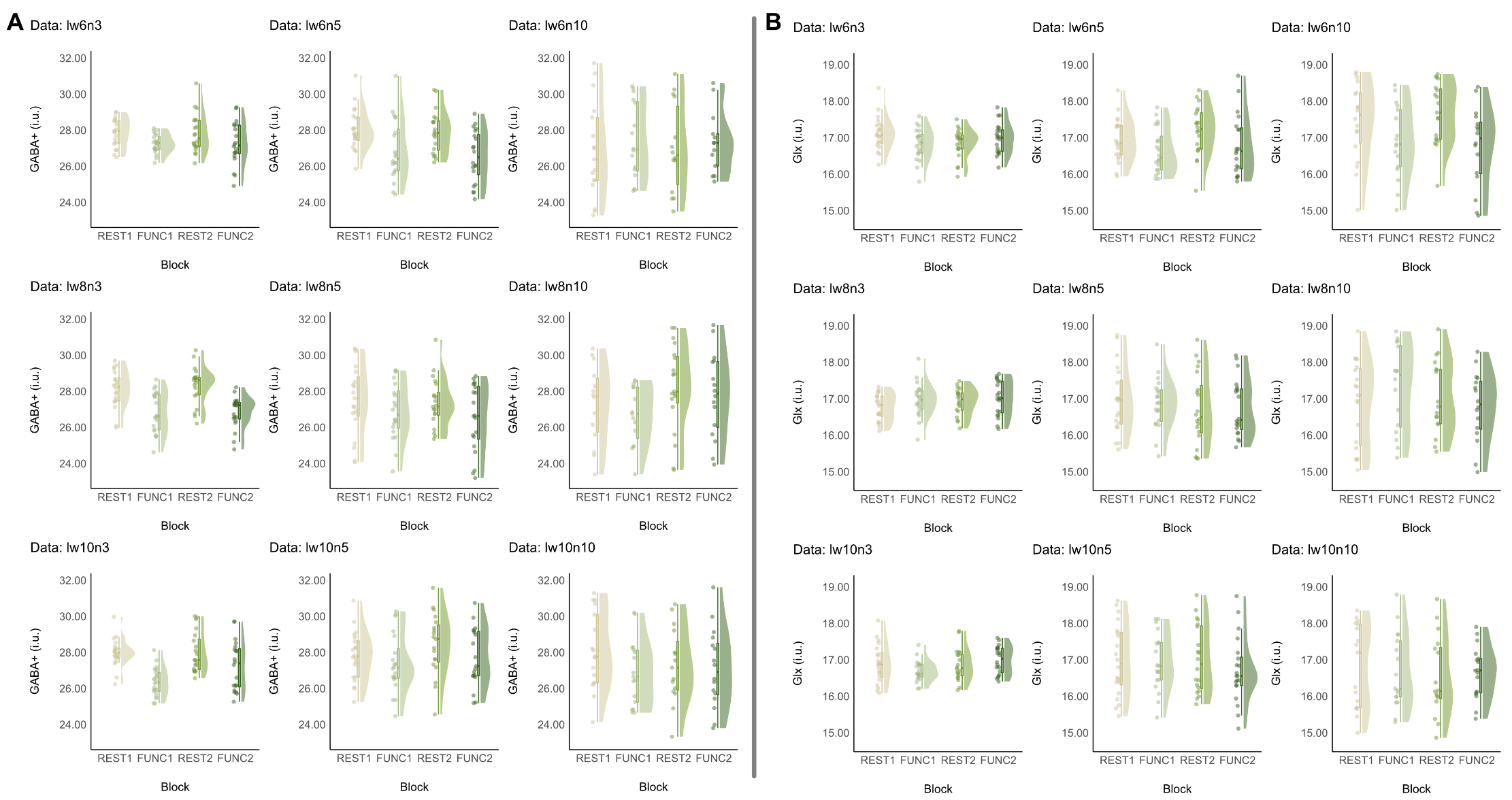

Supplementary Figure 3 (A) GABA+ (i.u.) and (B) Glx (i.u.) of simulated fMRS data acquired with block analysis for each data quality level. lw represents linewidth (Hz), and n represents noise_SD_ applied to the simulated dataset. The raincloud plot depicts individual data points, the half-violin plot represents data distribution, and the box plot with whiskers indicates the median and interquartile range. Data that fail quality metrics have not been plotted or included in the analysis.

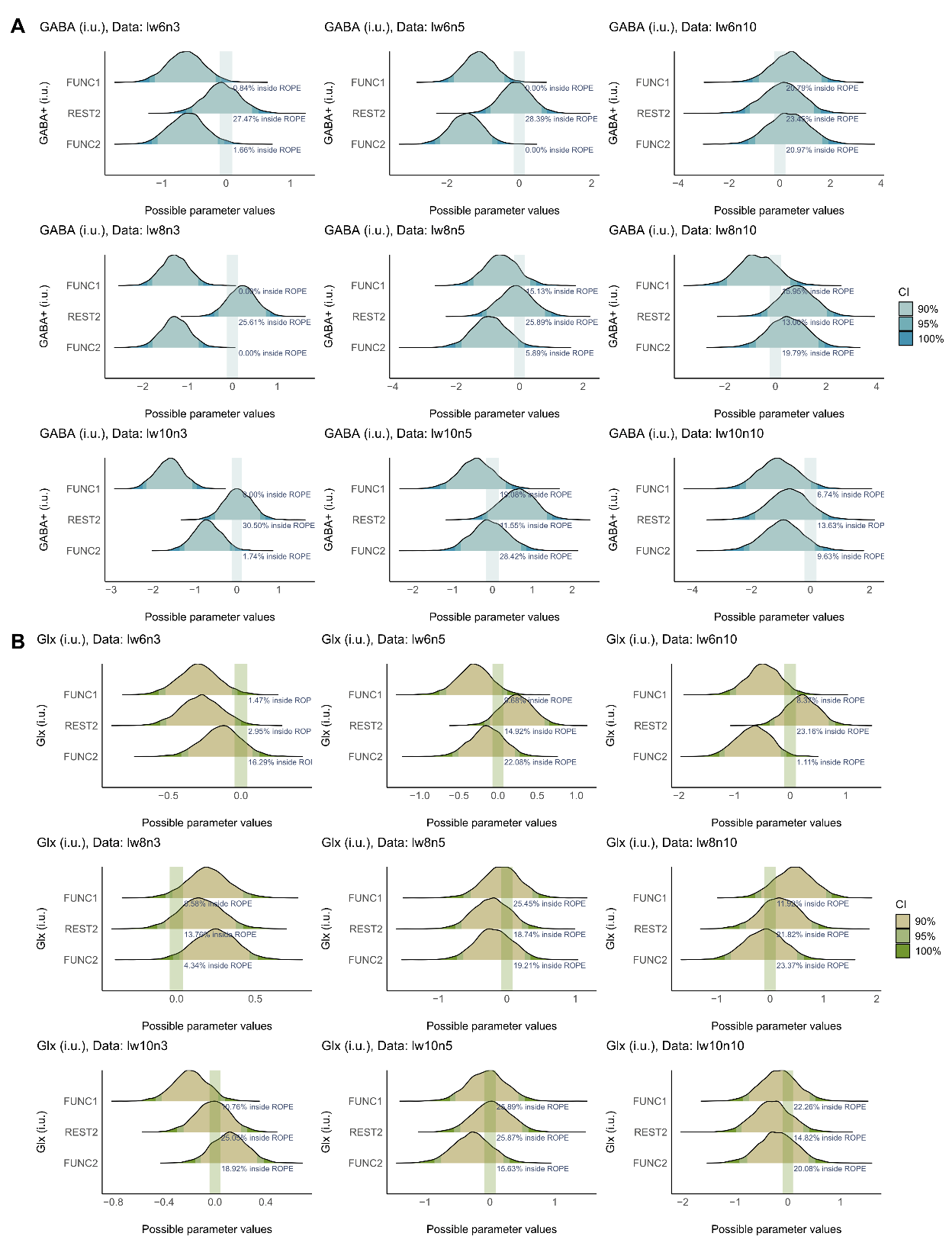

Supplementary Figure 4 Posterior distribution plots of (A) GABA+ (i.u.) and (B) Glx (i.u.) of simulated fMRS data analysed with block analysis for varying data quality. Different shades representing the range within the posterior distribution that contains a specified proportion (90%, 95% and 100%) of the probability density. The superimposed green boxes show ROPE region (-0.1 to 0.1 times the standard deviation of the outcome variable (J. K. Kruschke, 2018) that denotes the range considered practically equivalent to a negligible effect. The percentage displayed shows the portion of the 95% HDI of the posterior distributions that falls within the ROPE.

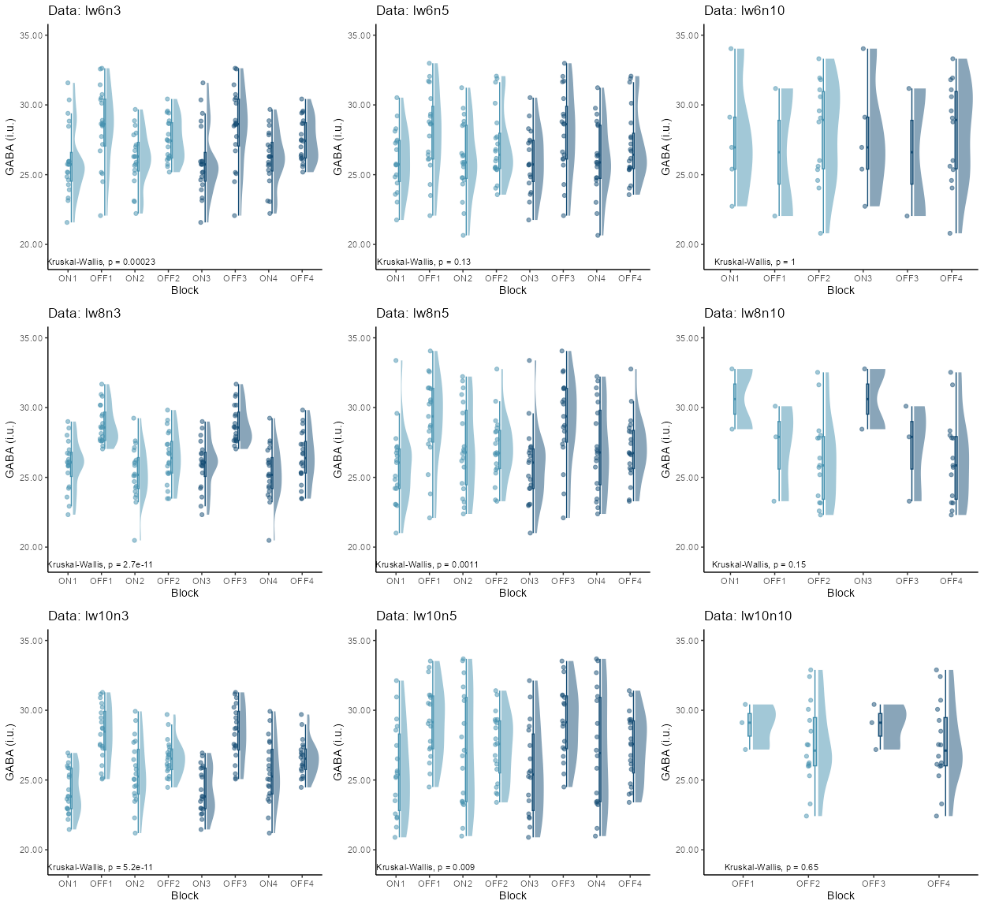

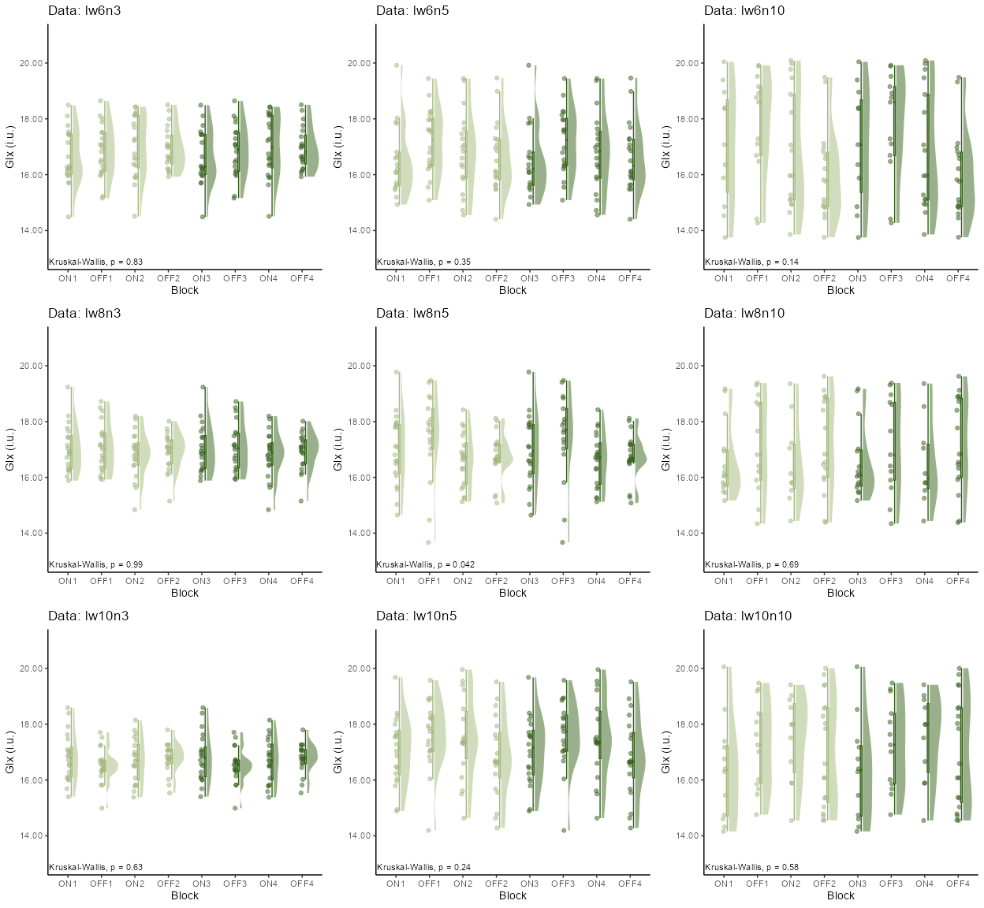

Supplementary Figure 5 (A) GABA+ (i.u.) and (B) Glx (i.u.) of simulated fMRS data obtained with event-related analysis based on the stimulus onset of 2s, the data were grouped in half to retain SNR for metabolite quantification. FUNC1 halved into ON1, ON2, OFF1, OFF2; FUNC2 halved into ON3, ON4, OFF3 and OFF. The raincloud plot depicts individual data points, the half-violin plot represents data distribution, and the box plot with whiskers indicates the median and interquartile range. Data that fail quality metrics have not been plotted or included in the analysis.

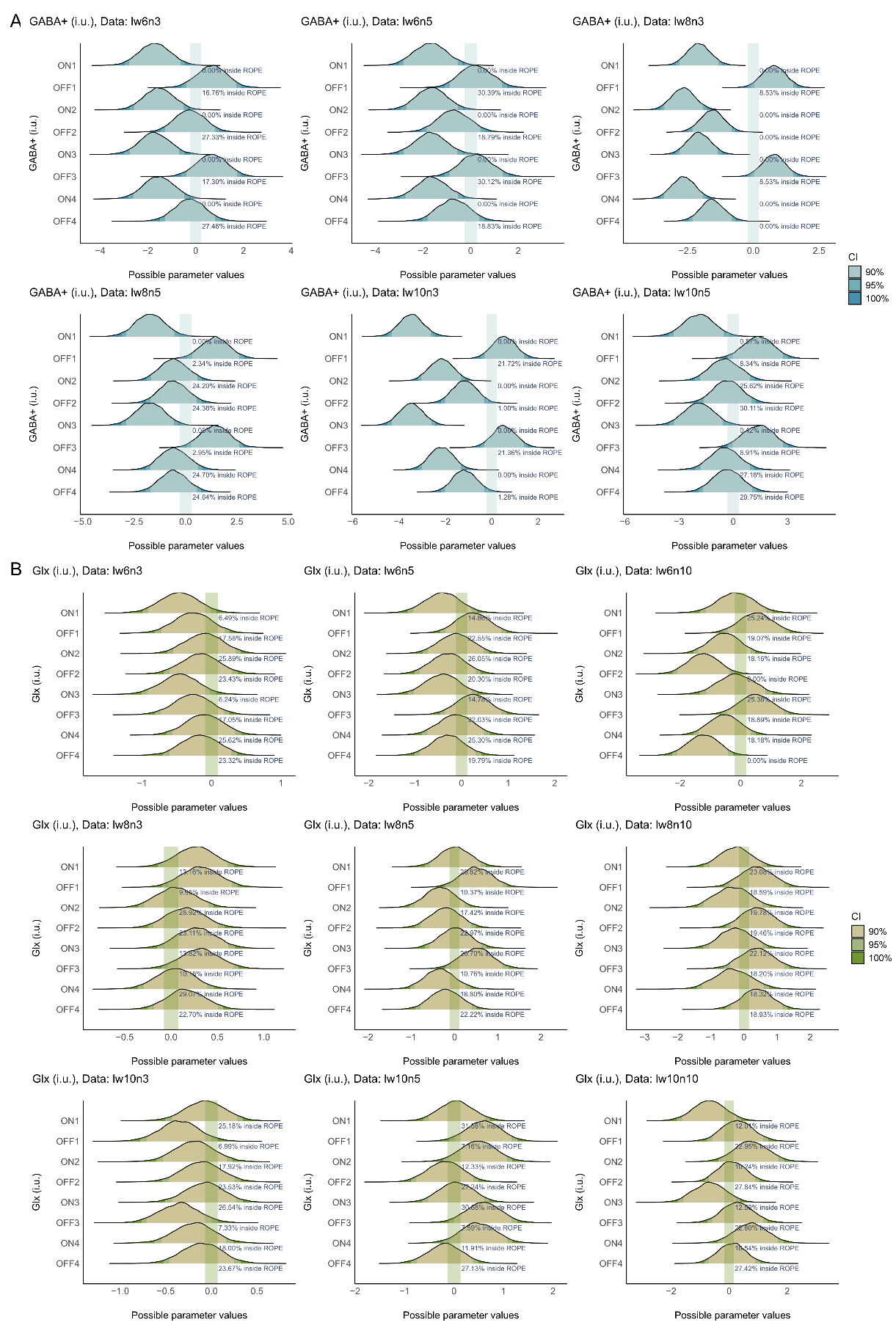

Supplementary Figure 6 ROPE analysis for (A) GABA+ (i.u.) and (B) Glx (i.u.) of simulated fMRS data obtained with event-related analysis. FUNC1 halved into ON1, ON2, OFF1, OFF2; FUNC2 halved into ON3, ON4, OFF3 and OFF4.

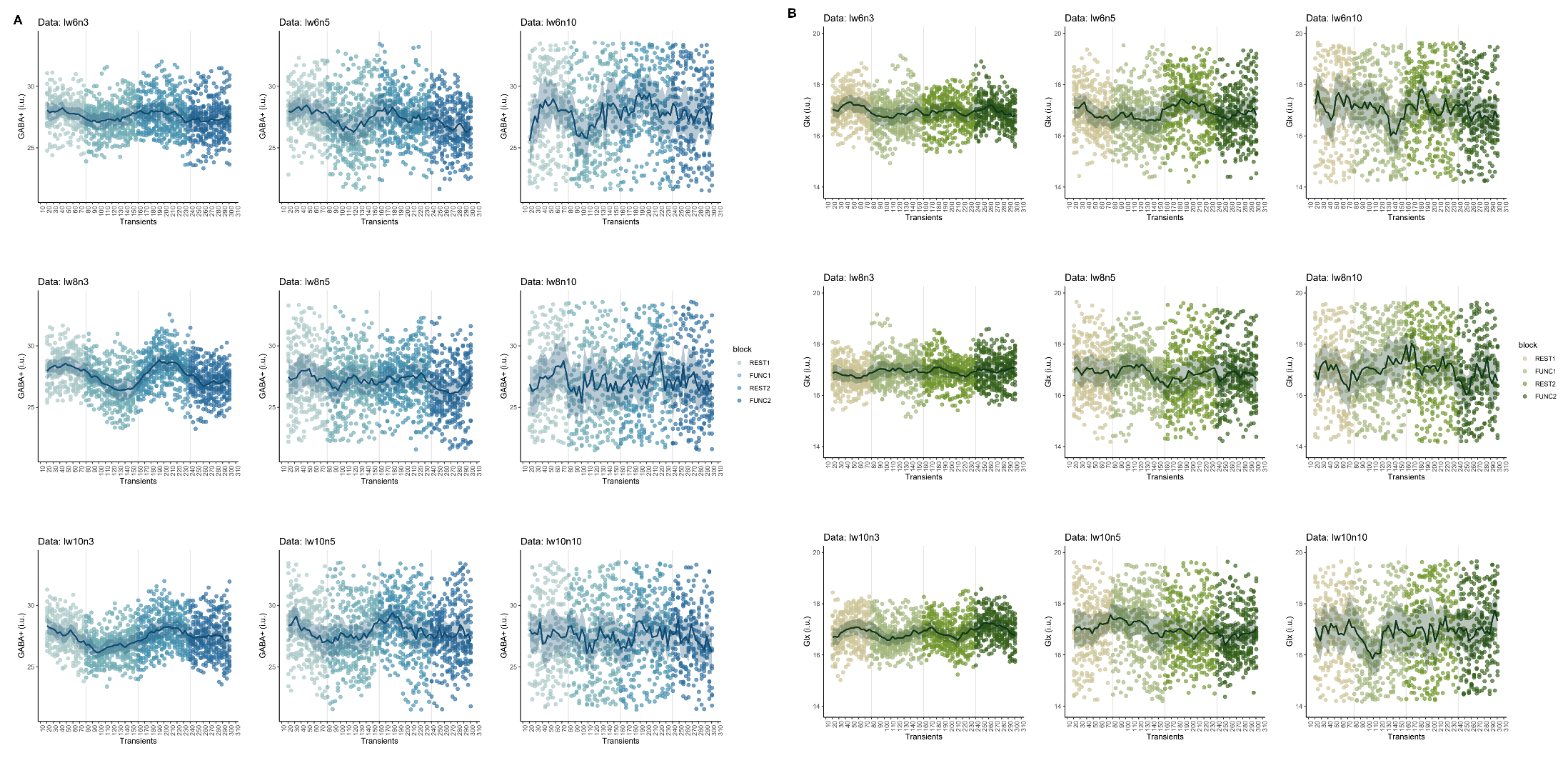

Supplementary Figure 7 Metabolite levels in i.u. unit of simulated fMRS data, analysed using sliding window analysis with varying transient sizes (ww). (A) GABA+(i.u.), (B) Glx(i.u.). The solid line represents the mean metabolite level, while the shaded area indicates the 95% confidence interval for each window.

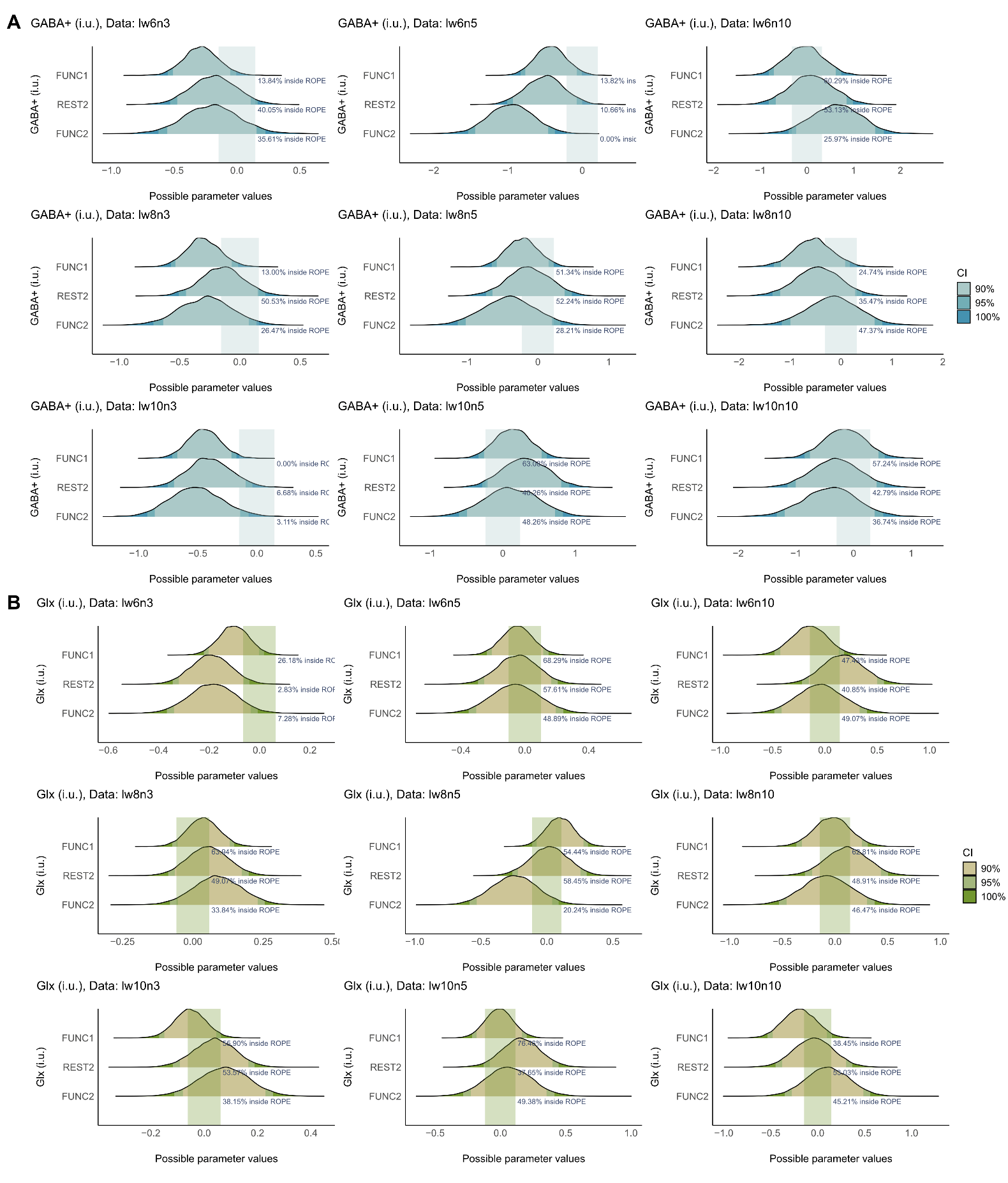

Supplementary Figure 8 ROPE analysis for (A) GABA+ (i.u.) and (B) Glx (i.u.) obtained using sliding window analysis showing multicollinearity (r> 0.7) between REST2 and FUNC2. For GABA+(i.u.), multicollimearity is observed in lw6n3 (n=0.71), lw6n5 (r=0.74), lw8n3 (r = 0.71), lw8n5 (r = 0.73), lw10n3 (r = 0.76), and lw10n5 (r = 0.79); for Glx(i.u.), multicollinearity is observed in lw6n5 (r=0.74), lw6n10 (r = 0.72), lw8n3 (r = 0.73), lw8n5 (r = 0.78), lw10n3 (r = 0.75), and lw10n5 (r = 0.79).

Supplementary Table 1 Average metabolite concentrations included in the simulated spectrum. Modified from (Govindaraju et al., 2000; Near et al., 2013).

| Metabolite | Concentration (mM) |
| --- | --- |
| Aspartate | 1.50 |
| Creatine | 5.25 |
| Phosphocreatine | 4.75 |
| Glycerophosphocholine | 1.00 |
| Phosphocholine | 0.60 |
| GABA | 1.90 |
| Glutamine | 3.20 |
| Glutamate | 9.50 |
| Glutathione | 1.50 |
| Myo-inositol | 6.00 |
| Lactate | 0.40 |
| N-Acetylaspartate | 12.00 |
| N-Acetylaspartylglutamate | 1.50 |
| Scyllo-inositol | 0.45 |

Supplementary Table 2 Linewidth and amplitude of simulated MM peaks relative to the signal of the one proton Cr peak. Modified from (Near et al., 2013; Oeltzschner et al., 2020).

| Macromolecule | Chemical shift | Linewidth (Hz) | Amplitude |
| --- | --- | --- | --- |
| MM09 | 0.91 | 21.00 | 3.00 |
| MM20 | 2.08 | 22.20 | 1.33 |
|  | 2.25 | 24.60 | 0.33 |
|  | 1.95 | 18.50 | 0.33 |
|  | 3.00 | 24.60 | 0.4 |
| MM12 | 1.21 | 24.60 | 2.00 |
| MM14 | 1.43 | 24.60 | 2.00 |
| MM17 | 1.67 | 21.00 | 0.20 |
| MM30 | 3.00 | 14.00 | 2.28 |

*Supplementary Table 3 Posterior distribution from Bayesian linear-mixed model for each model from block analyses for baseline MRS data. ^a,b^ Based on SEXIT framework, threshold for Significant (a) and large (b) effect is 0.05*SD_y_ and 0.3*SD_y_ where SD_y_ is the standard deviation of the outcome parameter (Makowski et al., 2019a).*

| Parameter | Median | 95% CI | pd | Significance^a^ | Large^b^ | Rhat | ESS |
| --- | --- | --- | --- | --- | --- | --- | --- |
| Glx/tCr |  |  |  | >\|1.04e-03\| | > \|6.26e-03\| |  |  |
| (Intercept) | 1.20E-01 | [ 0.12, 0.13] |  |  |  |  |  |
| 160 | -5.82E-05 | [-3.31E-03, 3.23E-03] | 51.28% | 0.28 | 0.01% | 1 | 29035 |
| 80 | -2.39E-04 | [-3.26E-03, 2.76E-03] | 56.20% | 0.30 | 5.00E-05% | 1 | 31946 |
| 64 | -1.37E-04 | [-3.05E-03, 2.78E-03] | 53.49% | 0.27 | 0% | 1 | 31096 |
| 32 | -5.00E-05 | [-2.90E-03, 2.77E-03] | 51.39% | 0.25 | 5.00E-05% | 1 | 33256 |
| 16 | 3.77E-04 | [-2.43E-03, 3.12E-03] | 60.57% | 0.32 | 0% | 1 | 32692 |
| GABA+/tCr | |  |  | >\|9.95e-04\| | >\|5.97e-03\| |  |  |
| (Intercept) | 0.12 | [0.12, 0.13] |  |  |  |  |  |
| 160 | -7.97E-05 | [-5.42e-03, 5.33e-03] | 51.24% | 0.37 | 1.54% | 1 | 36286 |
| 80 | 6.54E-04 | [-4.21e-03, 5.48e-03] | 60.02% | 0.44 | 1.53% | 1 | 40979 |
| 64 | 1.29E-04 | [-4.59e-03, 4.93e-03] | 52.18% | 0.36 | 0.91% | 1 | 43271 |
| 32 | 4.97E-04 | [-3.96e-03, 5.13e-03] | 58.87% | 0.42 | 1.04% | 1 | 46507 |
| 16 | 1.65E-03 | [-2.86e-03, 6.16e-03] | 76.13% | 0.61 | 2.97% | 1 | 46724 |
| Glx(i.u.) |  |  |  | > \|0.08\| | > \|0.50\| |  |  |
| (Intercept) | 8.09E+00 | [ 7.66, 8.52] |  |  |  |  |  |
| 160 | 2.00E-02 | [-0.21, 0.24] | 55.38% | 0.28 | 0% | 1 | 13496 |
| 80 | 2.00E-02 | [-0.19, 0.22] | 56.75% | 0.27 | 0% | 1 | 12751 |
| 64 | 2.00E-02 | [-0.18, 0.22] | 57.77% | 0.27 | 0% | 1 | 12550 |
| 32 | 4.00E-02 | [-0.15, 0.24] | 66.88% | 0.35 | 0% | 1 | 12129 |
| 16 | 8.00E-02 | [-0.11, 0.27] | 79.29% | 0.49 | 5.00E-05% | 1 | 11962 |
| GABA+(i.u.) | |  |  | > \|0.02\| | > \|0.13\| |  |  |
| (Intercept) | 2.22E+00 | [ 2.10, 2.33] |  |  |  |  |  |
| 160 | -5.94E-03 | [-0.11, 0.10] | 54.77% | 0.38 | 70.70% | 1 | 10461 |
| 80 | 1.00E-02 | [-0.08, 0.11] | 60.63% | 0.42 | 0.66% | 1 | 9542 |
| 64 | 2.00E-02 | [-0.07, 0.11] | 64.55% | 0.46 | 0.78% | 1 | 9414 |
| 32 | 1.00E-02 | [-0.07, 0.10] | 62.28% | 0.43 | 0.46% | 1 | 9436 |
| 16 | 4.00E-02 | [-0.04, 0.13] | 83.78% | 0.68 | 2.14% | 1 | 9174 |

*Supplementary Table 4 Posterior distribution from Bayesian linear-mixed model for each model from sliding window analyses of baseline MRS data. ^a,b^ Based on SEXIT framework, Threshold for Significant (a) and large (b) effect is 0.05*SD_y_ and 0.3*SD_y_ where SD_y_ is the standard deviation of the outcome parameter* (Makowski et al., 2019a)*.*

| Parameter | Median | 95% CI | pd | Significance^a^ | Large^b^ | Rhat | ESS |
| --- | --- | --- | --- | --- | --- | --- | --- |
| Glx/tCr |  |  |  | > \|1.17e-03\| | > \|7.01e-03\| |  |  |
| (Intercept) | 1.20E-01 | [0.12, 0.13] |  |  |  |  |  |
| 160 | 1.62E-03 | [-1.02e-03, 4.26e-03] | 88.06% | 0.63 | 0.01% | 1 | 29886 |
| 80 | 1.51E-03 | [-9.76e-04, 4.00e-03] | 88.45% | 0.61 | 0% | 1 | 39080 |
| 64 | 1.83E-03 | [-8.18e-04, 4.47e-03] | 91.01% | 0.69 | 0% | 1 | 38444 |
| 32 | 2.28E-03 | [-3.63e-04, 4.90e-03] | 95.26% | 0.79 | 0% | 1 | 38715 |
| 16 | 3.20E-03 | [5.27e-04, 5.83e-03] | 99.05% | 0.93 | 0% | 1 | 39626 |
| GABA+/tCr |  |  |  |  |  |  |  |
| (Intercept) | 0.12 | [0.12, 0.13] |  |  |  |  |  |
| 160 | 2.91E-03 | [-1.05e-03, 6.88e-03] | 92.45% | 0.92 | 83.27% | 1 | 24425 |
| 80 | 3.97E-03 | [3.20e-04, 7.72e-03] | 98.30% | 0.98 | 94.50% | 1 | 28053 |
| 64 | 4.76E-03 | [8.38e-04, 8.70e-03] | 99.08% | 0.99 | 97.16% | 1 | 26948 |
| 32 | 5.02E-03 | [1.02e-03, 9.00e-03] | 99.25% | 0.99 | 97.66% | 1 | 26831 |
| 16 | 8.00E-03 | [3.97e-03, 0.01] | 99.99% | 1 | 99.94% | 1 | 26696 |
| Glx(i.u.) |  |  |  | > \|0.06\| | > \|0.39\| |  |  |
| (Intercept) | 7.22E+00 | [6.83, 7.63] |  |  |  |  |  |
| 160 | 7.00E-02 | [-0.08, 0.22] | 83.23% | 0.55 | 0% | 1 | 15581 |
| 80 | 8.00E-02 | [-0.06, 0.22] | 87.08% | 0.59 | 0% | 1 | 12587 |
| 64 | 1.00E-01 | [-0.05, 0.25] | 89.85% | 0.67 | 0.01% | 1 | 12496 |
| 32 | 1.10E-01 | [-0.04, 0.26] | 92.47% | 0.72 | 0.02% | 1 | 12277 |
| 16 | 1.60E-01 | [6.71e-03, 0.31] | 97.94% | 0.89 | 0.18% | 1 | 12281 |
| GABA+(i.u.) |  |  |  | > \|0.02\| | > \|0.10\| |  |  |
| (Intercept) | 1.99E+00 | [ 1.90, 2.07] |  |  |  |  |  |
| 160 | 5.00E-02 | [-0.01, 0.11] | 93.27% | 0.84 | 6.29% | 1 | 8089 |
| 80 | 6.00E-02 | [ 0.00, 0.13] | 98.09% | 0.93 | 11.77% | 1.001 | 5586 |
| 64 | 7.00E-02 | [ 0.01, 0.14] | 98.41% | 0.95 | 18.08% | 1.001 | 5488 |
| 32 | 8.00E-02 | [ 0.01, 0.15] | 99.19% | 0.97 | 26.28% | 1.001 | 5405 |
| 16 | 1.10E-01 | [ 0.05, 0.18] | 99.98% | 1 | 64.89% | 1.001 | 5641 |

Supplementary Table 5 Posterior distribution from the Bayesian linear-mixed model of GABA+/tCr and GABA+ (i.u.) for each model from the block analyses for simulated fMRS data at each data quality. Only datasets with significant results (95% HDI falling outside the ROPE region) are shown. ^a,b^ Based on SEXIT framework, Threshold for Significant (a) and large (b) effect is 0.05*SD_y_ and 0.3*SD_y_ where SD_y_ is the standard deviation of the outcome parameter (Makowski et al., 2019a).

| Parameter  Data quality | Median | 95% CI | pd | Significance^a^ | Large^b^ | Rhat | ESS |
| --- | --- | --- | --- | --- | --- | --- | --- |
| lw6n5: GABA+/tCr |  |  |  | *> 4.06E-04* | *>0.00243* |  |  |
| (Intercept) |  |  |  |  |  |  |  |
| FUNC1 | -5.85E-03 | [-0.01, -1.22e-03] | 99.28% | 0.99 | 0.92 | 1 | 6692 |
| REST1 | -7.64E-04 | [-5.66e-03, 4.02e-03] | 62.60% | 0.56 | 0.25 | 1 | 5725 |
| FUNC2 | -7.77E-03 | [-0.01, -2.89e-03] | 99.90% | 1 | 0.98 | 1 | 5613 |
| lw8n3: GABA+/tCr |  |  |  | *> 0.000333* | *> 0.002* |  |  |
| (Intercept) |  |  |  |  |  |  |  |
| FUNC1 | -6.49E-03 | [-9.95e-03, -3.07e-03] | 99.98% | 1 | 1 | 0.999 | 5430 |
| REST1 | 8.69E-04 | [-2.58e-03, 4.19e-03] | 68.90% | 0.62 | 0.24 | 1 | 5531 |
| FUNC2 | -6.46E-03 | [-9.89e-03, -3.07e-03] | 100% | 1 | 0.99 | 1 | 5417 |
| Lw10n3: GABA+/tCr |  |  |  | *>3.31E-04* | *>1.99E-03* |  |  |
| (Intercept) |  |  |  |  |  |  |  |
| FUNC1 | -8.27E-03 | [-0.01, -4.78e-03] | 99.98% | 1 | 1 | 0.999 | 5620 |
| REST1 | 3.42E-05 | [-3.40e-03, 3.55e-03] | 50.70% | 0.44 | 0.14 | 1 | 5960 |
| FUNC2 | -3.35E-03 | [-6.85e-03, 7.38e-05] | 97.25% | 0.96 | 0.79 | 1 | 5459 |
| lw6n5: GABA+ (i.u.) |  |  |  | *>0.08* | *>0.45* |  |  |
| (Intercept) |  |  |  |  |  |  |  |
| FUNC1 | -1.11E+00 | [-1.96, -0.27] | 99.30% | 0.99 | 0.94 | 1.001 | 3853 |
| REST1 | -9.00E-02 | [-0.92, 0.77] | 58% | 0.52 | 0.2 | 1.001 | 3578 |
| FUNC2 | -1.46 | [-2.32, -0.59] | 99.88% | 1 | 0.99 | 1.002 | 4023 |
| lw6n5: GABA+ (i.u.) |  |  |  | >0.06 | >0.37 |  |  |
| (Intercept) |  |  |  |  |  |  |  |
| FUNC1 | -1.28E+00 | [-1.90, -0.65] | 100.00% | 1 | 1 | 1.001 | 3399 |
| REST1 | 2.10E-01 | [-0.40, 0.85] | 75% | 0.69 | 0.31 | 1.002 | 3040 |
| FUNC2 | -1.27E+00 | [-1.89, -0.66] | 100.00% | 1 | 1 | 1.001 | 3342 |
| lw6n5: GABA+ (i.u.) |  |  |  | *>0.06* | *>0.37* |  |  |
| (Intercept) |  |  |  |  |  |  |  |
| FUNC1 | -1.60E+00 | [-2.24, -0.95] | 100.00% | 1 | 1 | 1 | 3369 |
| REST1 | 2.00E-02 | [-0.63, 0.68] | 52% | 0.45 | 0.15 | 1 | 3800 |
| FUNC2 | -7.10E-01 | [-1.36, -0.05] | 98.20% | 0.97 | 0.84 | 1 | 3563 |
